# The use of a Cohort Size Shrinkage Index (CSSI) to quantify regional famine intensity during the Chinese famine of 1959-1961

**DOI:** 10.1101/2021.12.24.21268375

**Authors:** Chunyu Liu, Chihua Li, Hongwei Xu, Zhenwei Zhou, L.H. Lumey

**Affiliations:** Department of Epidemiology and Biostatistics, School of Public Health, Peking University, Beijing, China; Department of Epidemiology, Mailman School of Public Health, Columbia University, New York, NY, USA; Zhengzhou Central Hospital Affiliated to Zhengzhou University, Henan, China; Department of Sociology, Queens College, City University of New York, New York, NY, USA; Department of Biostatistics, Boston University School of Public Health, Boston, MA, USA

**Keywords:** Chinese, Regional famine intensity, Birth cohort trend, Dose-response, Tuberculosis after famine

## Abstract

The Cohort Size Shrinkage Index (CSSI) is the most widely used intensity measure in studies of the Chinese famine of 1959-1961. We examined requirements to construct a valid CSSI measure: reliable information on birth cohort size; a stable cohort size trend outside the famine years; and limited migrations. We used census data to examine the cohort size trend and concentrated on the time window of 1950-1970 to exclude events with a large impact on cohort size trends other than the famine. We observed a significant difference in cohort size trends comparing pre-famine and post-famine births. Compared to pre-famine based CSSI (p-CSSI), pre- & post-famine based CSSI (pp-CSSI) tends to overestimate famine intensity at higher mean CSSI levels and underestimate intensity at lower mean levels. In Sichuan province, we demonstrated a less pronounced dose-response relation between famine intensity and tuberculosis outcomes using p-CSSI as compared to pp-CSSI. These observations demonstrates that the CSSI is not a robust measure as had been assumed previously. We recommend the use of p-CSSI and encourage researchers to re-examine their results of Chinese famine studies.

## Introduction

The Great Chinese Famine of 1959-1961 (Chinese famine) originated from a combination of radical agricultural and economic policies and crop failures (Cao 2005; Chang and Wen 1998; Lin and Yang 1998, 2000; Yang 1996). The famine led to over 30 million excess deaths and is the largest famine recorded (Chang and Wen 1997; Garnaut 2014; Peng 1987; Zhao and Reimondos 2012). It had different intensity levels across regions (Ashton et al. 1992; Cao 2005; Chang and Wen 1997; Peng 1987). There has been an extensive literature on the famine’s demographic consequences (Aird 1982; Ashton et al. 1992; Banister 1984; Cai and Feng 2005; Coale and Banister 1994), its main causes (Chang and Wen 1998; Kasahara and Li 2020; Kung and Lin 2003; Meng, Qian and Yared 2015), and its potential long-term health and economic impact (Almond et al. 2007; Fan and Qian 2015; Li and Lumey 2017; Xu et al. 2016). Chinese famine studies rely heavily on appropriate indicators of famine intensity as these are being used either as dependent or as independent variables in statistical models to address specific research questions (Li and Lumey 2017; Li et al. 2019).

It has been a challenge to construct robust indicators of the famine’s intensity. First, there is no general agreement on what elements should be used to define and assess a famine’s intensity and magnitude (Gráda 2009; Howe and Devereux 2004). An optimal measure will depend on the field of research and specific research questions. Second, there is scant reliable information on food availability and consumption at the time of Chinese famine and mortality data are incomplete (Li and Lumey 2017; Li et al. 2019; Lumey, Stein and Susser 2011). Facing these challenges, scholars have proposed a number of demographic indicators to quantify regional famine intensity in China (Li 2022). These include the excess death rate (EDR) (Chang and Wen 1998; Peng 1987; Yang 1996), the cohort size shrinkage index (CSSI) (Huang et al. 2010b; Xu et al. 2016), the abnormal death proportion (Cao 2005; Song, Wang and Hu 2009), and the infant mortality rate (Song et al. 2009). Although none of the indicators are direct measures of famine intensity - as this would require recording individual level calorie intakes, they do reflect demographic changes caused by a famine and capture important aspects of a famine’s intensity.

Among famine intensity measures, the EDR and CSSI have traditionally been the most widely used. In recent years, the CSSI has enjoyed a growing popularity as an alternative to the EDR, especially in economics and the health sciences (**Figure 1**). This is because of concerns about the quality of mortality statistics at the time of the famine and their limited accessibility at the local prefecture and county levels (Huang et al. 2010b; Xu et al. 2016). CSSIs avoid these potential problems as they are derived from population census data that are deemed to be more reliable. CSSIs have therefore been increasingly applied to relate famine intensity to later economic and health outcomes in populations affected by famine (**Table A1 of the online Appendix**).

**Fig. 1.**
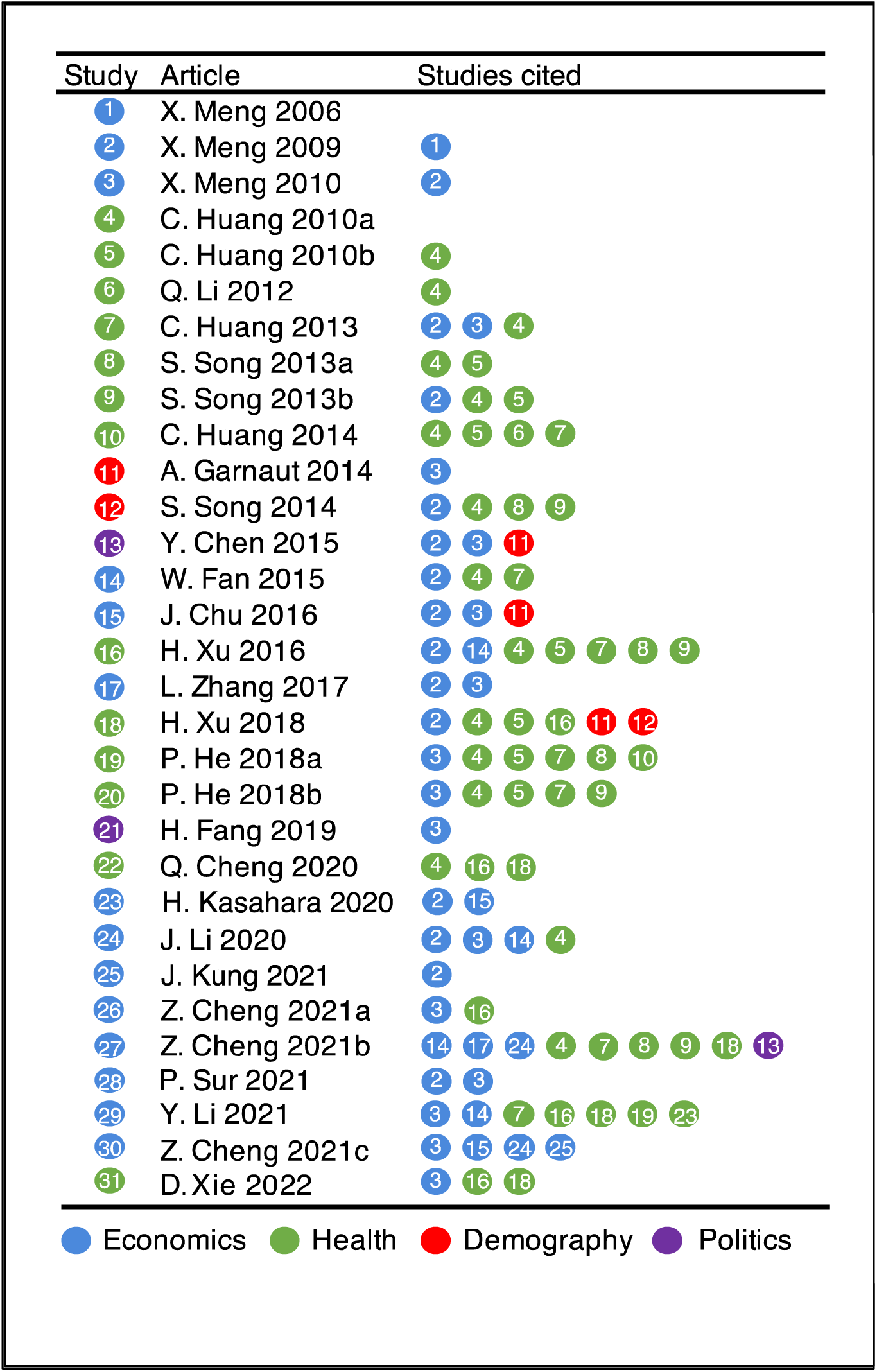
Chinese famine studies using cohort size shrinkage index (CSSI). Detailed information of listed studies is available in Table A1.

The CSSI compares observed cohort sizes of populations born in famine years to expected cohort sizes in the absence of famine and is estimated by averaging cohort sizes of pre- & post-famine births (Huang et al. 2010b; Xu et al. 2016). There has been little consistency, however, in exactly how CSSIs were constructed or applied in different studies. There has been a wide variety in the selection of data sources, estimating methods, geographic regions, and definitions of famine and non-famine years (**Table A1**). Almost all studies combined pre- & post-famine births to estimate an expected cohort size for famine years (**Figure A1**). As we later demonstrate, CSSIs estimated from pre- & post-famine births can introduce unrecognized biases when estimating a potential dose-response relationship between famine intensity and later health outcomes when pre-famine and post-famine cohort trends are different(Li, Zhou and Lumey 2021).

For CSSI to be a useful measure of famine intensity at the local level, reliable information will be needed on several variables: first on the size of birth cohorts, perhaps from census data at the province or prefecture level; second, to confirm a stable cohort size trend over time in the non-famine period as this will be needed to identify any famine related deviations from existing trends, and third, on migrations between localities if place of birth is not known and populations have to be classified instead by current residence, an imperfect indicator of place of birth (Xu et al. 2016; Xu et al. 2018). To date, no studies have examined the adequacy of the information used to construct CSSIs. Also, it remains unknown how sensitive CSSI measures are to variations in their component parts. These variations include data sources and estimating methods.

In this study, we use the 1% China 2000 Census as the main data source to examine cohort sizes by year of birth. We construct CSSIs using varying combinations of non-famine years (pre-famine years vs pre- & post-famine years) and then compare CSSI distributions at the province and prefecture levels. We examine the impact of migrations on CSSI by comparing place of birth and residence from census information. We use the tuberculosis surveillance study in Sichuan province as a case-study to examine how empirical results may change with the application of CSSIs using non-famine years as reference (Cheng et al. 2021; Cheng et al. 2020; Li et al. 2021). Our findings demonstrate that the choice of non-famine years is by far the most important determinant of CSSI, shaping both its distribution and the results of empirical studies.

## Data and Methods

### Main Data and Variables

To date, cohort size shrinkage indexes (CSSIs) at different regional levels have been used by over 30 Chinese famine studies, using different national census waves collected between 1982-2000. (**Table A1**). This study is mainly based on the 1% China 2000 Census (https://international.ipums.org/international/) (Minnesota Population Center 2020), because it provides the most extensive demographic information, including birth year, place of birth at the province level, and place of residence both at the prefecture and province levels (Lavely 2001). This enables comparison of place of birth and place of residence to examine inter-province migrations. The 1% China 1982 and 1990 census do not provide information on place of birth but will be important for sensitivity analysis as described later.

Our study includes 29 province level regions and 340 prefectures. The province level regions are: Sichuan (SC), Anhui (AH), Guizhou (GZ), Hunan (HuN), Henan (HeN), Qinghai (QH), Jiangsu (JS), Ningxia (NX), Guangxi (GX), Gansu (GS), Shandong (SD), Yunan (YN), Hubei (HuB), Fujian (FJ), Hebei (HeB), Zhejiang (ZJ), Jiangxi (JX), Guangdong (GD), Liaoning (LN), Shanxi (SX), Shaanxi (SaX), Jilin (JL), Inner Mongolia (IM), and Heilongjiang (HLJ). Before the 1980s, Hainan was part of Guangdong province, and Chongqing (CQ) was part of Sichuan province. Separately grouped are the province level regions formed by the cities of Beijing (BJ), Tianjin (TJ) and Shanghai (SH) where population policies were different than in other regions and early family planning policies were introduced in the 1960s (Yang 2004). Also grouped apart are the two provinces Xinjiang (XJ) and Tibet (TB) to which government population policies encouraged active immigration (Sabit and Mamat 2008). We arranged the above listing of the 24 regular provinces by CSSI rank from the highest to the lowest and separately show the three city provinces and the two active immigration provinces (see also **Table A3**).

### Cohort Size Projection and CSSI Construction

The 2000 census includes births between 1900 and 2000. For optimal CSSI estimates, a relatively stable period for births will be required to use as a reference period against which to identify deviations caused by the famine. We selected the period 1950-1970 as the most suited for this purpose because its start marks the establishment of the People’s Republic of China in 1949 after chaotic periods of the Second Sino-Japanese War and the Chinese Civil War (Coale and Chen 1987; Xizhe 1989). And its end marks the introduction of family planning policies across China in the 1970s (Poston 1992). This window also includes the famine and non-famine years selected by previous Chinese famine studies that used CSSIs (**Figure A1**).

These famine studies calculated CSSI as 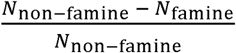 at different regional levels, where *N*_*famine*_ is the observed average cohort size of famine births and *N*_*non-famine*_ is the average cohort size of the combined pre- & post-famine births. The *N*_*non-famine*_ is assumed to reflect the expected cohort size in the absence of famine. However, the potential impact of different trends of pre-famine and post-famine births on CSSI has not been systematically examined before. Trends could be different because post-famine births in China show a rebound after the famine and this rebound could be related to the severity of the decline (Li et al. 2021). Therefore, CSSIs may also be sensitive to the choice of non-famine years (pre-famine years vs. pre- & post-famine years combined).

To examine these questions, we divided the period of 1950-1970 into three periods and defined 1950-1957 as pre-famine years, 1959-1961 as famine years, and 1963-1970 as post-famine years. We excluded births in 1958 or 1962 from analysis because births in these years could be misclassified on famine exposure status comparing different provinces with different famine start and end years. We examined birth trends based on pre-famine births and post-famine births alone and compared trends at the regional level.

We compared two approaches to estimating birth deficits during the famine period. The first examines deviations from expectations based on a linear extrapolation of pre-famine births (1950-1957) and the second examines deviations from the commonly used linear interpolation of combined pre- & post-famine births (1950-1957 & 1963-1970). This corresponds to calculating CSSIs from either pre-famine birth cohorts alone (p-CSSI) or combining pre- & post-famine birth cohorts together (pp-CSSI). Both p-CSSI and pp-CSSI were mapped at the province and prefecture levels. In sensitivity analyses, we compared findings based on the 1% China 1990 population census and examined the robustness of CSSI measures to variations in the exact years defined as non-famine years.

### Applying CSSIs to Tuberculosis Surveillance Study in the Sichuan Province

We applied the p-CSSI and pp-CSSI approaches to the empirical study in the Sichuan province of tuberculosis cases diagnosed between 2005 and 2018 (Cheng et al. 2020). The study data were publicly available at the Github repository https://github.com/qu-cheng/TB_famine. In line with previous approaches, we evaluated associations between famine intensity and tuberculosis risk by mixed-effects meta-regressions. (Cheng et al. 2021; Cheng et al. 2020; Li et al. 2021). For famine births, we fit linear models to examine the relation between famine intensity as represented by CSSI on the log of the ratio of the observed vs expected non-famine incidence rates. We modeled multiple sets of CSSI using either varying combinations of non-famine years (pre-famine years vs pre- & post-famine years) at the prefecture level in Sichuan province or the full Sichuan 2000 Census or the 1% China 2000 Census for Sichuan province.

## Results

### Cohort Size Trends Differ When Selecting Either Pre-Famine or Post-Famine Births as the Non-Famine Population

Panel a of **Figure 2** shows the observed cohort sizes by birth year between 1900 and 2000 at the national level as documented by the 1% China 2000 Census. For individuals born until the late 1950s, a monotone relation is seen between higher mortality and advancing age, demonstrating the smallest number of survivors among the oldest individuals. The trend is interrupted in three periods that show a marked decline in cohort size: for individuals born between 1958-1962, 1971-1980, and after 1990. A similar pattern is seen at the province level (**Figure A2**).

**Fig. 2.**
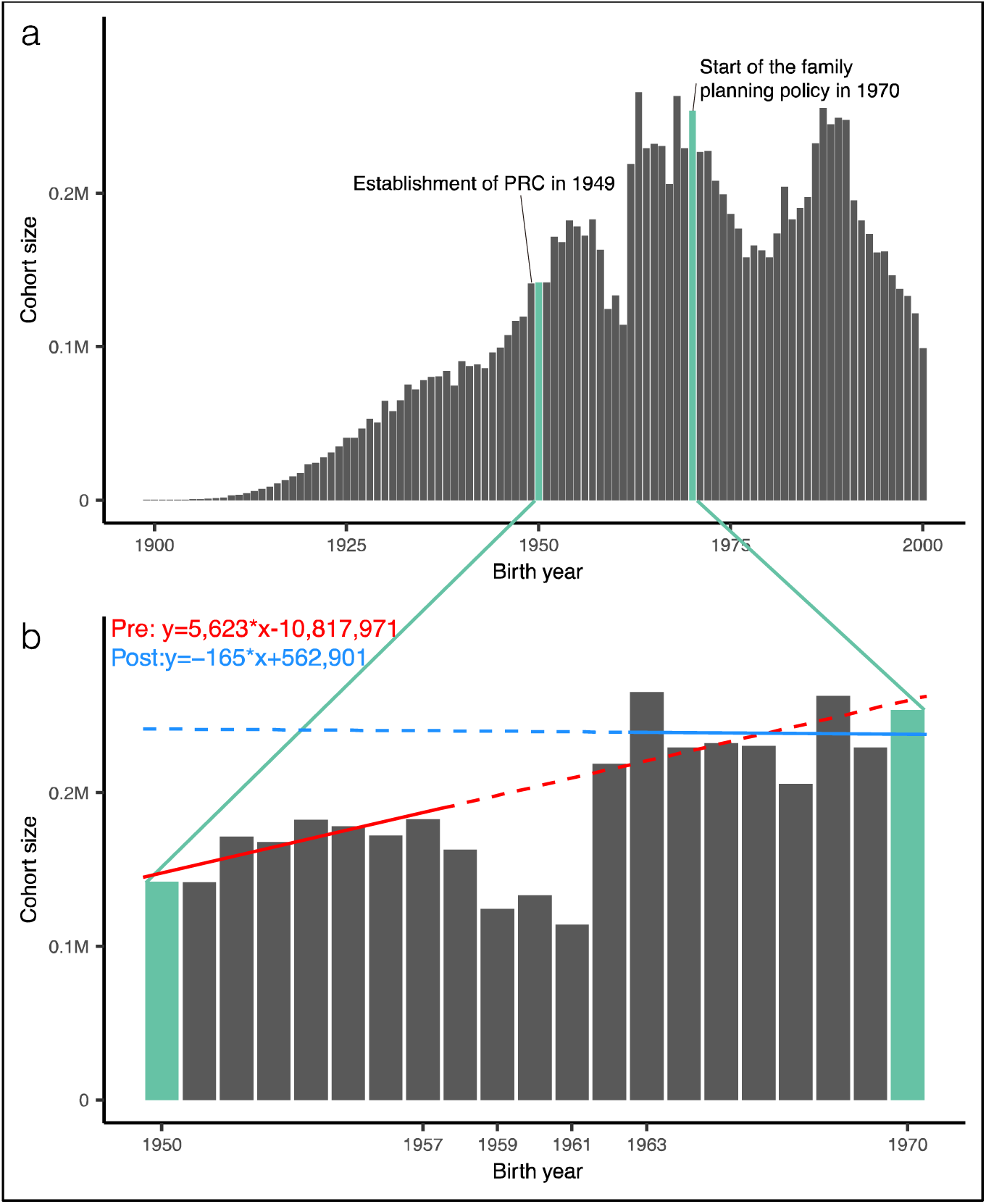
Cohort size by birth year in China. *Notes:* a. Cohort size by birth year, 1990-2000. b. Cohort size trend from linear regression either based on either pre-famine births (1950-1957) or post-famine births (1963-1970). *Source:* 1% China 2000 Census, place of birth.

Panel b of **Figure 2** shows the cohort size pattern between 1950 and 1970 and compares the national cohort size trend among pre-famine births (1950-1957) (Red hatched line) to post-famine births (1963-1970) (Blue hatched line). The first trend increases by birth year while the second is flat. At the province level, the pre-famine based cohort size trends are also different from the post-famine based trends (**Figure A3**). These differences remain after excluding the birth peak in 1963 immediately after the famine ended.

### CSSI Estimates Also Critically Depend on the Choice of Non-Famine Years

Because cohort size trends differ for pre-famine births and post-famine births, estimated CSSIs will likely vary depending on the choice of non-famine births. We compared the observed cohort sizes of famine births in 1959-1961 with the projected populations in these years estimated from linear regressions at the province level using either the pre-famine births (1950-1957) alone or the pre- & post-famine births (1950-1957 & 1963-1970) combined (**Table A2**). The projected populations are larger than the observed populations in all provinces. The estimated birth deficits also vary with the choice of non-famine years: estimated CSSIs based on pre-famine births as non-famine years (p-CSSI) differ from CSSIs based on pre- & post-famine births as non-famine years (pp-CSSI). **Table A3** shows the CSSI values and rankings for all provinces. The difference between p-CSSI and pp-CSSI ranges from -16.2% in Qinghai province (35.4% vs 51.6%) to +14.1% in Liaoning province (38.0% vs 23.9%). It is more extreme for the five regions grouped separately, ranging from -60.6% in Xinjiang (−38.8% vs 21.8%) to +39.3% in Shanghai (38.5% vs -0.8%) (**Table A4**).

The geographical distribution of CSSIs is shown in **Figure 3**, with p-CSSI in Panel a and pp-CSSI in Panel b. The Sichuan, Anhui, Guizhou and Hunan provinces show the largest estimated shrinkage regardless of estimation method. pp-CSSIs tend to be higher than p-CSSIs in the Midwest of China however and lower in the Northeast and Southeast of China (Panel c). Compared to p-CSSIs, pp-CSSIs tend to be systematically larger at higher CSSI values and smaller at lower values with a crossover at the mean (Panel d). Although p-CSSI and pp-CSSI have similar mean values (37.4% vs 37.7%), their distributions differ, the former values being spread more narrowly around the mean (SD 9.8) than the latter (SD 14.5) (Appendix **Figure A4)**.

**Fig. 3.**
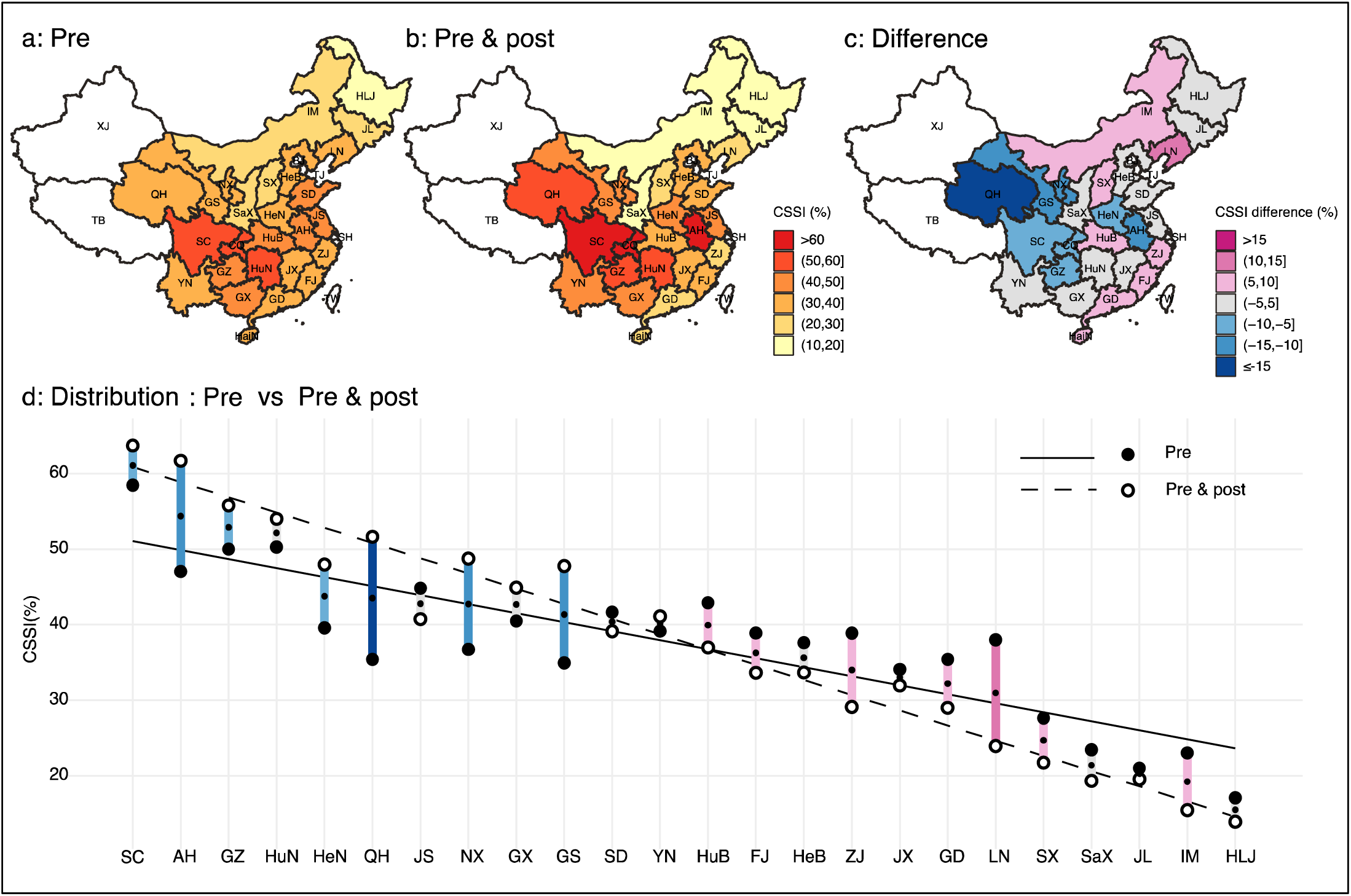
P-CSSI, pp-CSSI, and their difference at the province level. *Notes:* a. Provincial p-CSSI (%) based on births in 1950-57. b. Provincial pp-CSSI (%) based on births in 1950-57 & 1963-70. c. Difference of provincial CSSI (%) between panel a and panel b. d. Provinces are ordered by the average of p-CSSIs and pp-CSSIs. The solid line represents the linear regression of p-CSSI (filled circles) over the average of the two CSSIs. The dash line represents the linear regression of pp-CSSI (unfilled circles) over the average of two CSSIs. Each bar represents the difference of p-CSSIs and pp-CSSIs for each province, and its color coding follows the panel c. Full names corresponding to province abbreviations can be found in ‘Data and Methods’ section. *Source:* 1% China 2000 Census, place of birth.

### Other Factors Besides the Choice of Non-Famine Years Have Little Effect on CSSI

First, our findings do not depend on the choice of census waves. The 1% China 1990 Census has been the most widely used data source for previous studies although place of birth was not available (**Table A1**). CSSIs as estimated from the 1% 1990 census show the same cross-over pattern **(Figure A5)** as the 1% 2000 census. Second, we compared p-CSSI using place of residence from the 1990 or the 2000 census and place of birth from the 2000 census. The findings are highly consistent with inter-correlations exceeding 0.95 (**Table A5**). Third, p-CSSI estimates are robust to adding a quadratic term to linear regression trend extrapolations; fourth, to changing pre-famine years from 1950-1957 to 1940-1957; and fifth, to limiting analysis to the rural population instead of the entire population, with inter-correlations ranging from 0.95 to 0.99 for all comparisons.

### P-CSSI And Pp-CSSI are not Only Different at the Provincial but Also at the Prefecture Level

Because information on place of birth at the prefecture level was not available in any of the census waves, CSSI at this level was constructed instead using place of residence. **Figure 4** shows spatial variations of residence based CSSIs at both the province and prefecture levels. Panels a to c of **Figure 4** are comparable to those of **Figure 3**, showing findings at the province level using either information on place of birth or place of residence. Panels d and e of **Figure 4** show substantial variations at the prefecture level within most provinces. This is illustrated numerically by the wide range of p-CSSI at the prefecture level (**Table A6**). Panel f of **Figure 4** shows that the differences between p-CSSI and pp-CSSI at the province level also hold at the prefecture level. In the online Appendix, **Table A7** presents p-CSSIs and pp-CSSIs at the prefecture level for use in future comparative studies.

**Fig. 4.**
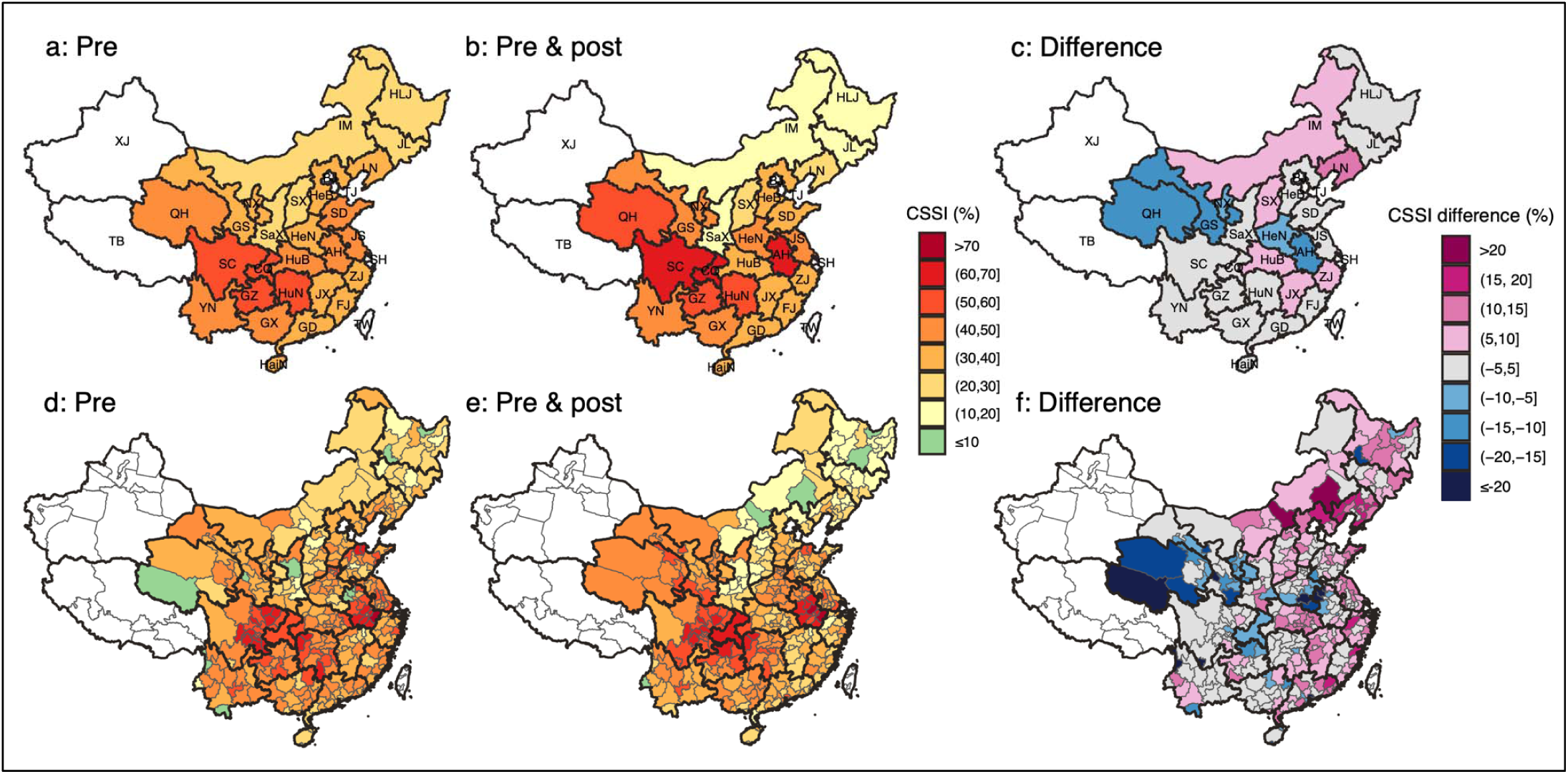
P-CSSI, pp-CSSI, and their difference at the province level (a-c) and prefecture level (d-f). *Notes:* a and d: Provincial and prefecture p-CSSI (%) based on births in 1950-57. b and e: Provincial and prefecture pp-CSSI (%) based on births in 1950-57 & 1963-70. c: Difference of provincial CSSI (%) between a and b. f: Difference of prefecture CSSI (%) between d and e. *Source:* 1% China 2000 Census, place of residence.

### Differences in P-CSSI and Pp-CSSI Characteristics Generate a Different Dose-Response Pattern Comparing Famine Intensity to Adult Health

A recent study in Sichuan province used the pp-CSSI to report a dose-response relationship between famine intensity and adulthood tuberculosis at the prefecture level (Cheng et al. 2020). We compared p-CSSI and pp-CSSI in this study. p-CSSI had a smaller mean and wider range compared to pp-CSSI (**Table A8**). Using the reported full 2000 Sichuan Census (Cheng et al. 2021), the dose-response estimate for (log) tuberculosis IRR in relation to a CSSI unit increase was less than half for p-CSSI compared to pp-CSSI (IRR 0.38; 95% CI: -0.12, 0.88 vs IRR 0.91; 95% CI: 0.19, 1.62) (**Figure 5 and Table A9**). By bootstrapping, we estimated a significant IRR difference between the two estimates (0.55; 95% CI: 0.06, 10.4). This demonstrates how sensitive CSSIs can be to the choice of non-famine years and their effect on estimated relations between famine intensity and later health outcomes.

**Fig. 5.**
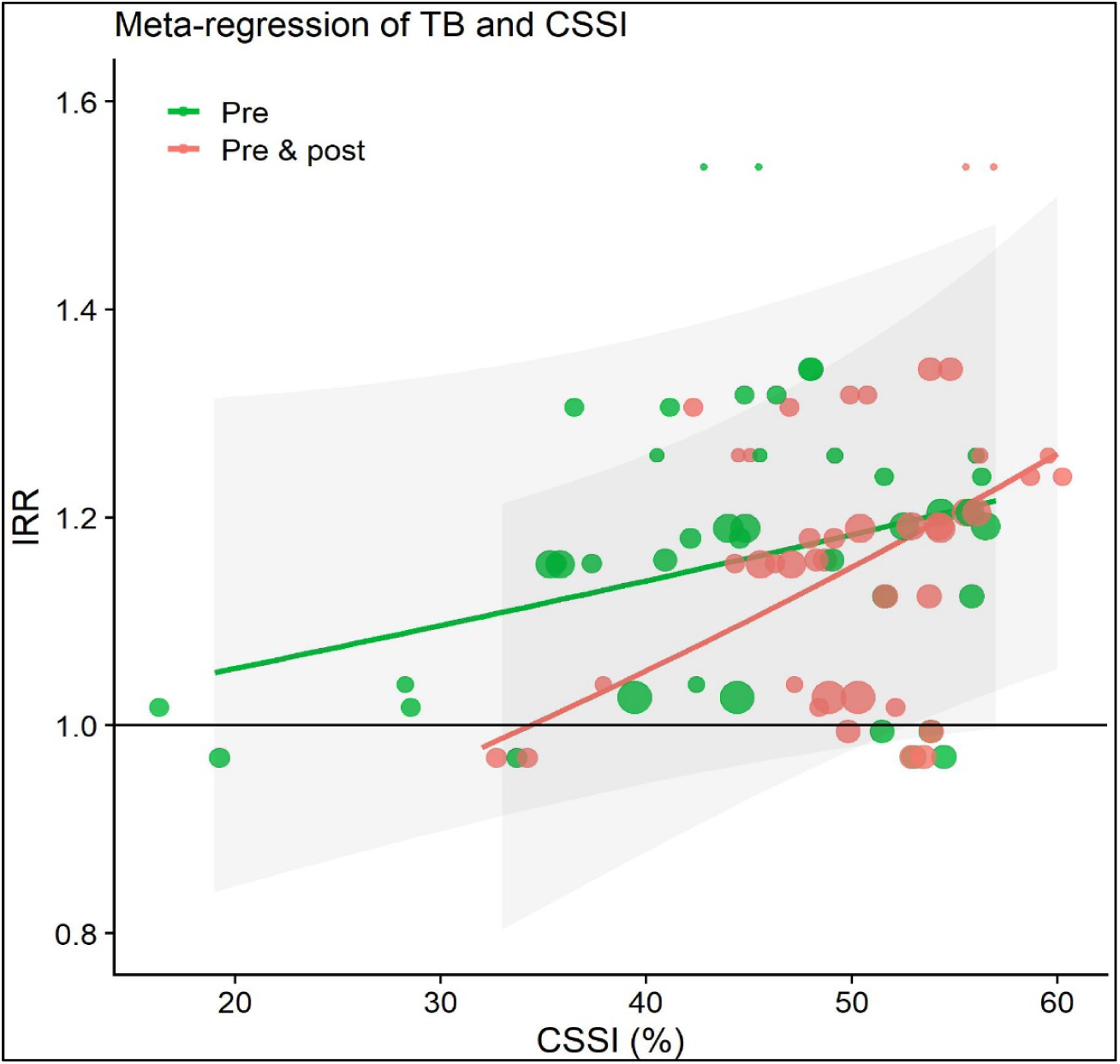
Scatterplot of CSSI and incidence rate ratio (IRR) of tuberculosis among F1 across prefectures in Sichuan province *Notes:* Each prefecture is represented by a dot. The size of the dot is proportional to the inverse variance of the estimated IRR of each prefecture. The lines represent the meta-regression fits, and the shaded areas represent the 95% CIs. *Source:* Full Sichuan 2000 Census, age at the time of the census and place of residence.

The p-CSSI based IRR is not sensitive to changes in the time window used to define pre-famine years, with an IRR of 0.37 (95% CI: -0.12, 0.86) using 1949-57 as pre-famine years and 0.40 (95% CI: -0.02, 0.82) using 1945-57 as pre-famine years compared to 0.38 (as in the above) for years 1950-1957. Findings using the 1% China 2000 Census data for Sichuan agree with these results from the full Sichuan 2000 Census.

## Discussion

In this study, we examined alternative approaches to construct CSSIs in Chinese famine studies and reviewed study findings based on this measure. We established that CSSI distributions can be highly sensitive to the choice of non-famine years (pre-famine years vs. pre- & post-famine years combined). This demonstrates that CSSI is not a robust estimator of famine intensity. By contrast, the choice of census waves, the use of province of residence vs province of birth, the exact timing of pre-famine years, and the form of statistical extrapolation models had little impact on CSSI. Perhaps of greater importance, we demonstrated that different CSSIs (p-CSSI vs. pp-CSSI) will generate a different understanding of the relation between famine intensity and later life health outcomes.

For Chinese famine studies, it is critically important to select time windows that are as free as possible from extraneous events (Garnaut 2014; Peng 1987; Zhao and Reimondos 2012). As an example of later post-famine events, family planning pilot programs introduced in some regions from the 1960s and nationwide in the 1970s most likely contributed to the cohort size trend irregularities observed by others for individuals (Aird 1982; Attane 2002; Poston Jr and Gu 1987). We took these circumstances into account by examining large cities in separate strata and limiting our post-famine observations to births 1963-1970.

Chinese famine studies to date have also failed to recognize that the cohort size trends among pre-famine births and post-famine births differ too much for pp-CSSI to be representative of a general trend. That this leads to biases can be seen from the comparison of pp-CSSI with p-CSSI measures that show systematic differences over the CSSI range: pp-CSSI tends to be systematically larger than p-CSSI at higher means of CSSI and lower at lower means of CSSI. A high correlation between declines in famine births and post-famine rebounds was already reported before (Huang et al. 2014; Huang et al. 2010a; Huang et al. 2010b; Huang et al. 2013). Our study confirms these observations based on a correlation of -0.36 and this finding was one of our motivations to recommend against the use of pp-CSSI. Other famine studies that also avoided pp-CSSI include the examination of excess deaths in China (Peng 1987) and in Ukraine for the 1932-1933 famine (Rudnytskyi et al. 2015).

Other factors besides including post-famine births had a limited impact on CSSI estimates. First, on the use of the 1% China 2000 Census as our main data source, we note that our study findings are highly consistent across census waves (**Table A5**). In different census waves, we found the similar systematic differences between p-CSSI and pp-CSSI. Second, migration could be an important factor influencing famine intensity measures, including CSSI. Using the 1% China 2000 Census however we found that place of birth was highly comparable to place of residence for estimating CSSI at the province level (**Figure A6**). This is consistent with our finding that inter-province migration among births 1950-1970 was as low as 6%. A recent study also showed that migration within provinces is not associated with famine intensity (Kasahara and Li 2020). Third, despite the substantial rural-urban difference in famine intensity because government policies prioritized food security in urban areas (Garnaut 2014; Peng 1987; Zhao and Reimondos 2012), our CSSI findings are not affected by ignoring urban populations. This can be explained by the fact that over 80 percent of Chinese population at the time lived in rural areas (Gørgens, Meng and Vaithianathan 2012; Peng 1987). Forth, linear and higher order statistical models and varying pre-famine years to project trends over time give the same results.

As a consequence, including post-famine births to estimating CSSIs has important consequences for understanding the relation between famine intensity experienced at the time of birth and health and economic outcomes in later life. In many current Chinese famine studies (**Table A1**), CSSI has been incorporated in regression models as a continuous measure of famine intensity. Changing cohort size trends of pre-famine births and post-famine births may not only affect CSSI estimates but also their distributions and the interpretation of study findings. We demonstrated that at the province level, pp-CSSI spreads more widely around the mean than p-CSSI. Using data from the tuberculosis surveillance study in Sichuan Province (Cheng et al. 2020), we demonstrated that the dose-response relation between famine intensity and tuberculosis appears to be much weaker using p-CSSI than using pp-CSSI, irrespective of the type of census data used. Our findings demonstrate in many ways how the choice of non-famine years determines the nature of observed dose-response relationships between famine intensity at the time of birth and adult health and their interpretation.

### Strengths and Limitations

Our study has many strengths. We use census data to construct robust measures of famine intensity and demonstrate that combining pre- & post-famine births can lead to unrecognized bias in estimating famine intensity using CSSI. In a study of later life health in Sichuan province, we show that combining pre- & post-famine births can amplify potential dose-response relations between famine exposure level and tuberculosis health outcomes. These strengths provide the basis for our recommendations for the future analysis of famine studies in China.

There are also some limitations. There is no gold standard for famine intensity. CSSI is a good indicator however of famine related fertility changes and excess deaths; Census data could be biased by selective survival after famine exposure in early life. But in the one study we know on the question (Song, 2009) this does not seem to be the case and our findings based on census waves ten years apart are highly consistent; Migrations can potentially lead to biases but had little impact at the province level. Further studies will be needed on migration at the prefecture level; Potential misclassifications of place of birth could arise if pregnant women migrated during the famine and gave birth in another provinces. A child’s place of birth was registered by mother’s Hukou status however which neutralizes this concern (Zhu 2010).

### Suggestions for Future Studies

We advocate the use of p-CSSI in Chinese famine studies to avoid biases related to different pre-famine and post-famine cohort size trends. Current studies (Cheng et al., 2020; Huang et al., 2014; Cheng Huang et al., 2010; Meng et al., 2015; Xu et al., 2016; Xu et al., 2018) should be re-analyzed with this concern in mind and divergent findings compared and discussed. For future studies, we provide data of p-CSSI and pp-CSSI at both the province and prefecture level in supplementary materials (**Table A7 of the online Appendix**). This information will be helpful in identifying provinces and prefectures across China where famine intensity was highest as these will be the most suitable locations for future famine studies. We first suggest studies in one or two of the most famine-severe provinces, including Sichuan and Anhui provinces (**Figure 4 & Table A7 of the online Appendix**). Prefecture level studies could be attempted later, guided by substantial variations in famine intensity within province. These studies will generate improved estimates of famine intensity at the local level as improve estimates of the nature of dose-response relationships between famine exposure and its long-term impact.

## Conclusions

Chinese famine studies to date have failed to recognize that cohort size trends among pre-famine births and post-famine births can differ substantially. This may be problematic as valid CSSI measures assume stable cohort size trends. We demonstrate for China that pre- & post-famine based CSSI is not a robust estimator of famine intensity as had been assumed previously. It will overestimate famine intensity at higher exposure levels and underestimate intensity at lower levels. As an alternative for Chinese famine studies, we therefore recommend CSSI based on pre-famine births alone.

## Data Availability

All data produced in the present work are contained in the manuscript.

https://github.com/chunyu-yes/CSSI_famine_severity

## Data Availability

All data and code necessary to reproduce the main findings of this study are available at the Github repository: https://github.com/chunyu-yes/CSSI_famine_severity.

## Acknowledgements

The authors are particularly grateful to the Minnesota Population Center for providing the data of the 1% China 1990 and 2000 Census, the National Bureau of Statistics in China for originally producing the census data, and the research team at Berkely University for making the data of tuberculosis surveillance study in the Sichuan province publicly available.

## Author contributions

C.L., C.L., and L.H.L designed research; C.L., C.L., H.X., Z.Z. and L.H.L performed research; C.L., C.L., H.X., Z.Z. and L.H.L analyzed data; C.L., C.L., and L.H.L drafted the paper; H.X. and Z.Z. provided technical support; and C.L., C.L., H.X., Z.Z. and L.H.L revised the paper.

