## Supplementary tables and figures for "The use of a Cohort Size Shrinkage Index (CSSI) to quantify regional famine intensity during the Chinese famine of 1959-1961"

Department of Epidemiology

Mailman School of Public Health

Columbia University

722 W 168th St, 1617A

New York, NY 10032.

**This file includes:**

Figures A1 to A6

Tables A1 to A9

SI References

### Supplementary figures:

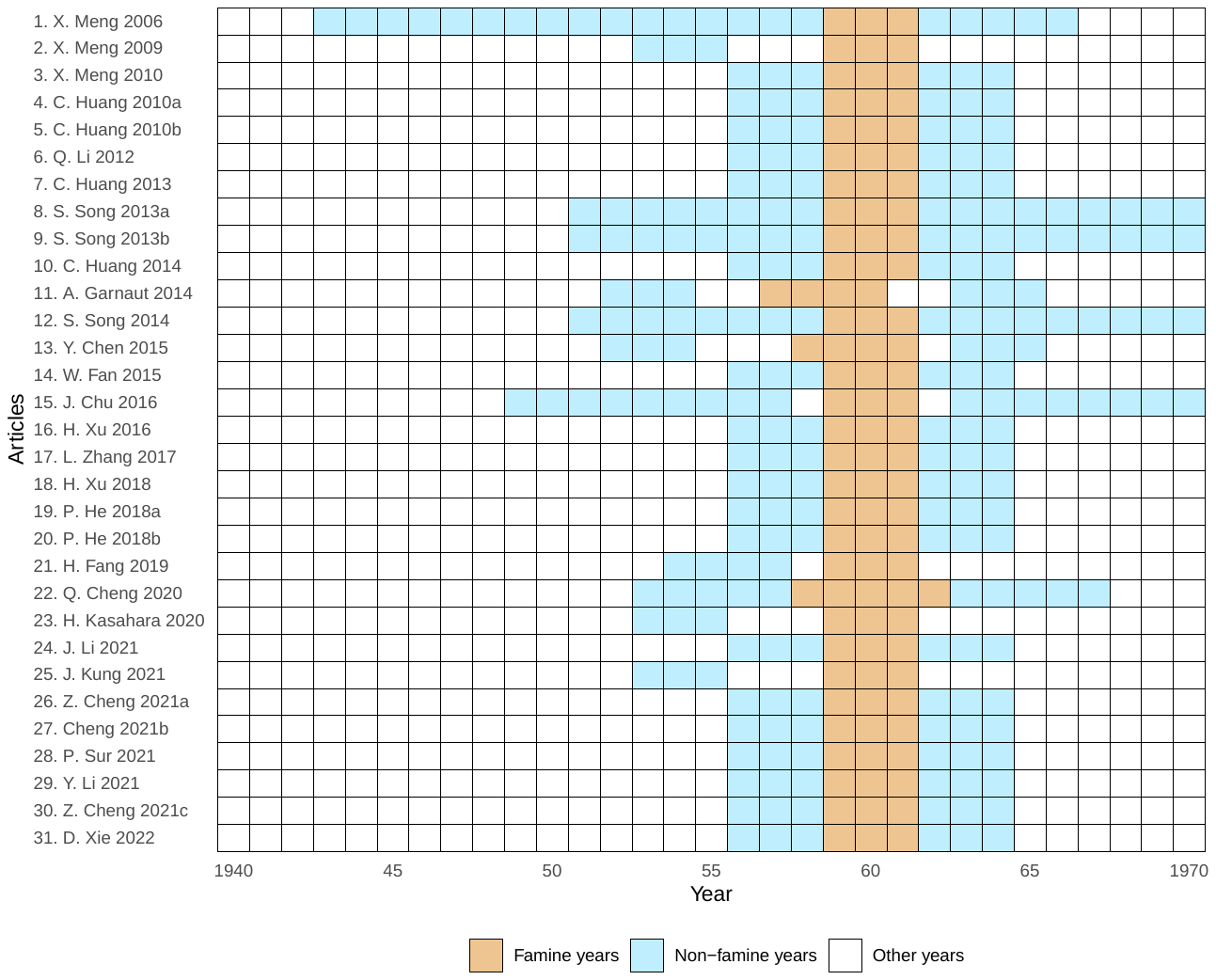

#### Fig A1. Famine and non-famine years as selected by Chinese famine studies using cohort size shrinkage index (CSSI).

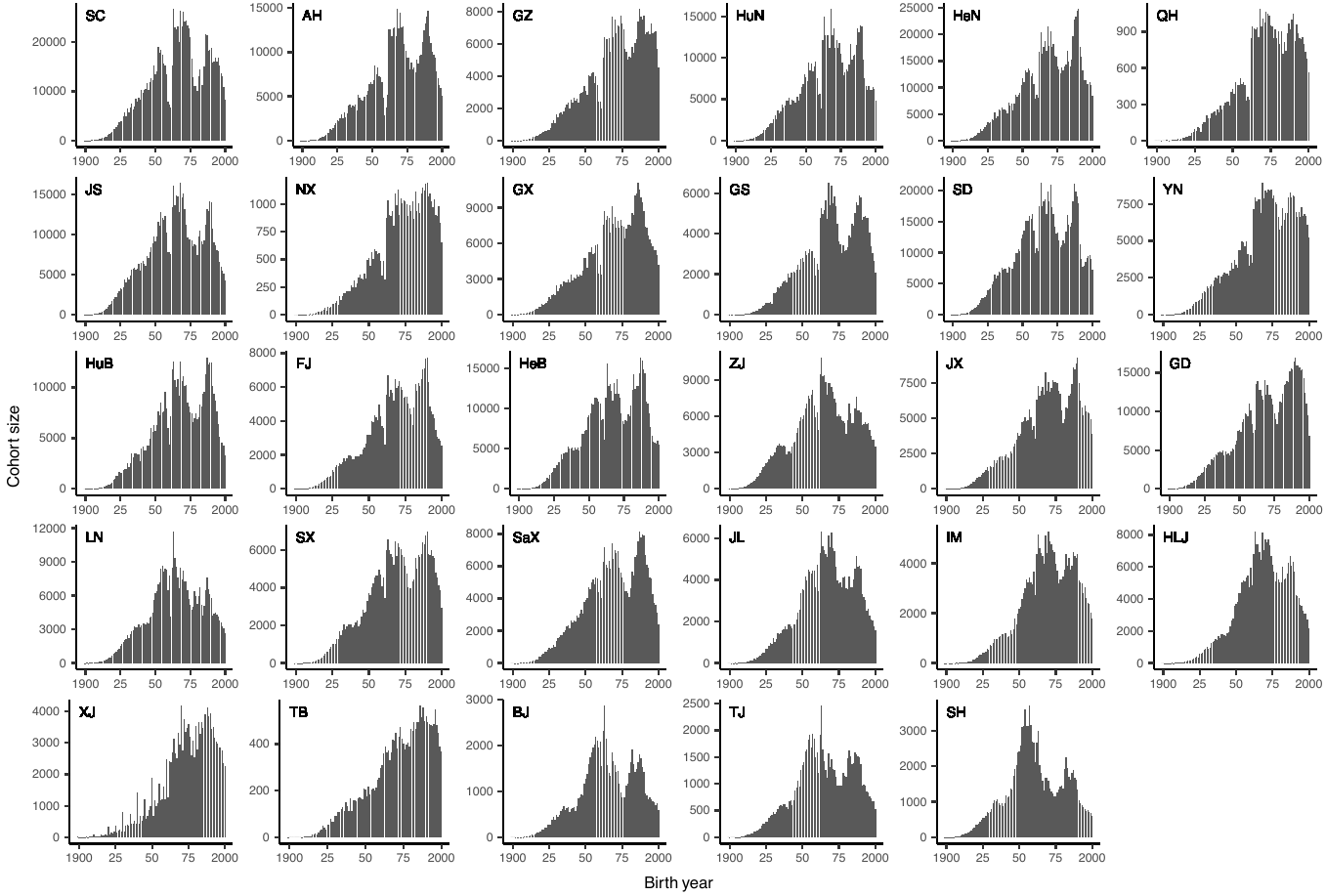

#### Fig A2. Cohort size by birth year, between 1900-2000 at the province level.

*Notes:* Full names corresponding to province abbreviations can be found in ‘Data and Methods’ section.

Y-axis scaled to population size in each province.

*Source:* 1% China 2000 Census, place of birth.

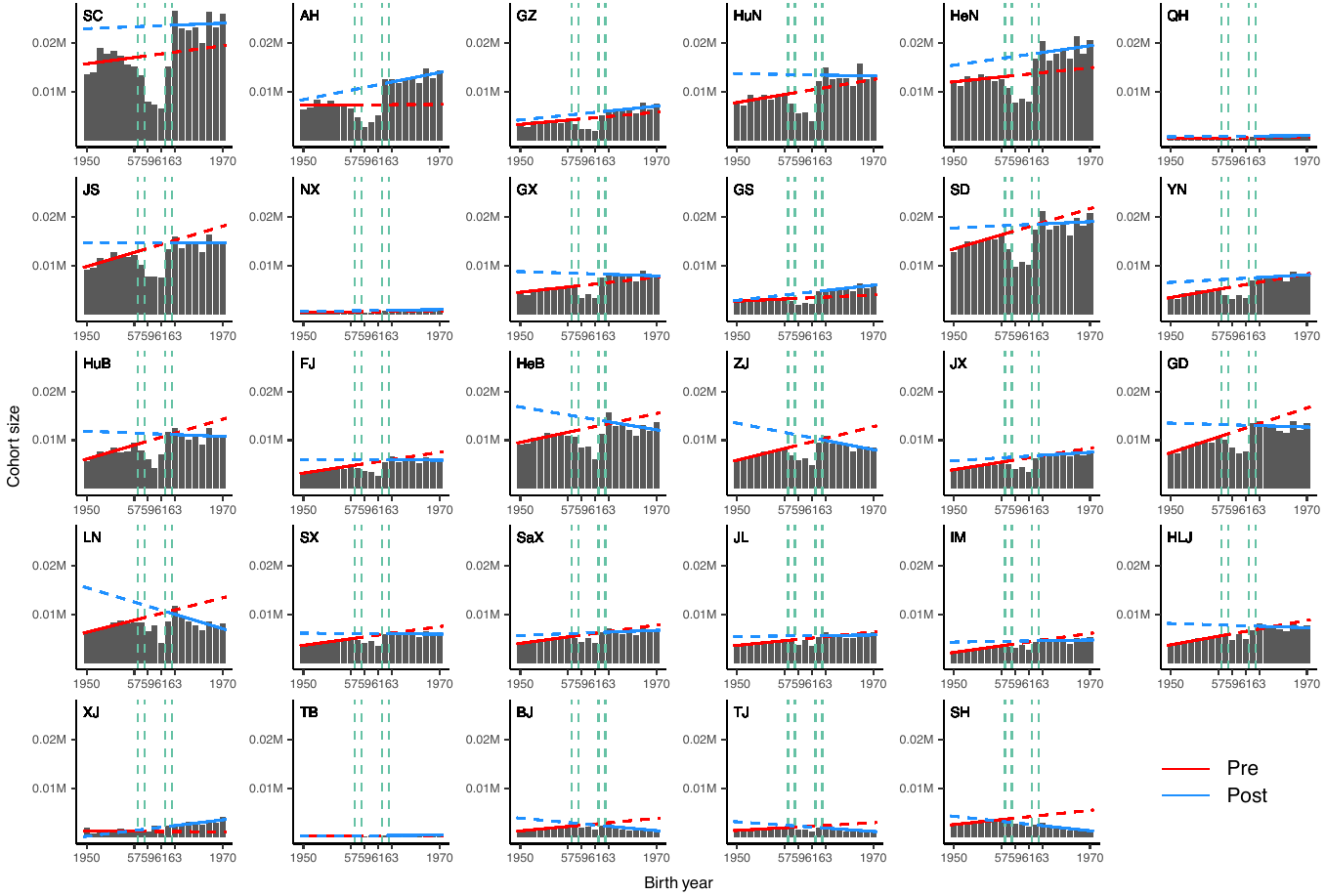

#### Fig A3. Cohort size trend from linear regression either based on either pre-famine births (1950-1957) or post-famine births (1963-1970) at the province level.

*Notes:* Full names corresponding to province abbreviations can be found in ‘Data and Methods’ section.

Y-axis not scaled to population size in each province.

*Source:* 1% China 2000 Census, place of birth.

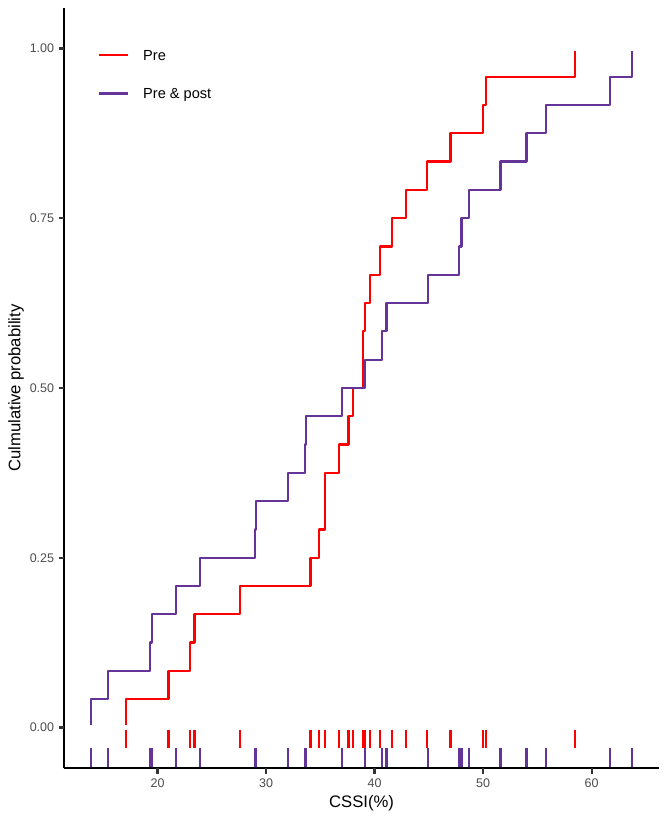

#### Fig A4. The distribution of p-CSSIs and pp-CSSIs at the province level.

*Notes:* This figure is produced based on data presented in Table S3.

*Source:* 1% China 2000 Census, place of birth.

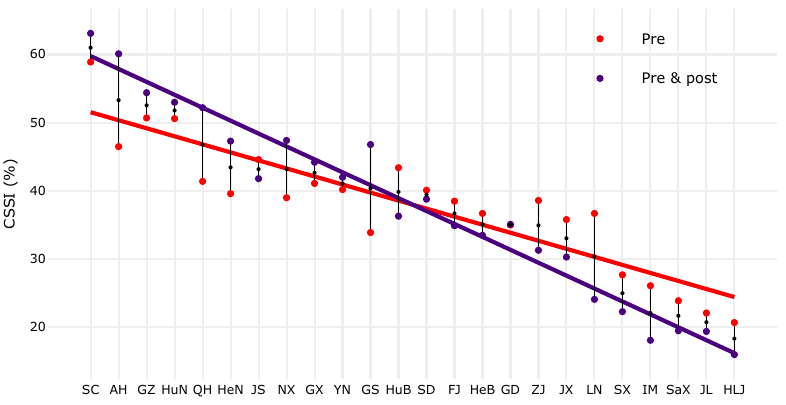

#### Fig A5. P-CSSIs and pp-CSSIs at the province level using 1% China 1990 Census.

*Notes:* Provinces are ordered by the average of CSSI based on pre-famine births and CSSI based pre- and post-famine births. The red line represents the linear regression of CSSI based on pre-famine births over the average of two CSSIs. The purple line represents the linear regression of CSSI based on pre- and post-famine births combined over the average of two CSSIs.

*Source:* 1% China 1990 Census, place of residence.

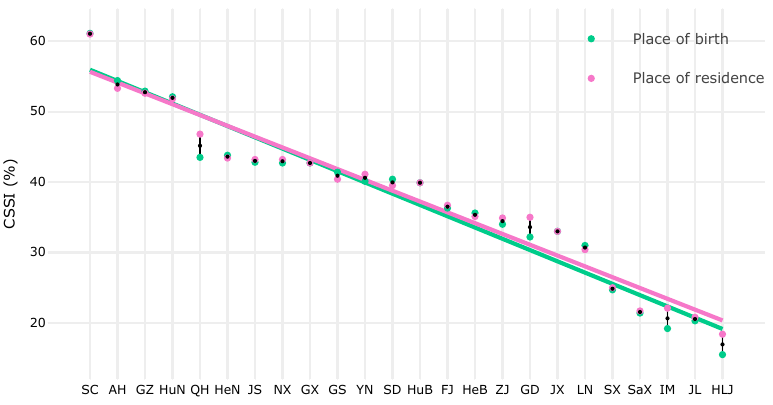

#### Fig A6. P-CSSIs at the province level, comparing place of birth and place of residence.

*Notes:* Provinces are ordered by the average of p-CSSIs based on place of birth and p-CSSIs based on place of residence. The green line represents the linear regression of p-CSSI based on place of birth over the average of two p-CSSIs. The pink line represents the linear regression of p-CSSI based on place of residence over the average of two p-CSSIs.

*Source:* 1% China 2000 Census, place of birth and residence.

### Supplementary Tables:

#### Table A1. Cohort size shrinkage index (CSSI) calculating characteristics in selected Chinese famine studies.

| Article | Data source | Estimating Method | Administrative  Level | Famine years | Non-famine years |
| --- | --- | --- | --- | --- | --- |
| 1. X. Meng 2006^(1)^ | 1% China 1990 census | Average cohort size | County level | 1959-61 | 1943-58 & 1962-66 |
| 2. X. Meng 2009^(2)^;  23. H. Kasahara 2020^(3)^;  25. J. Kung 2021^(4)^ | 1% China 1990 census | Average cohort size | County level | 1959-61 | 1952-54 |
| 3. X. Meng 2010^(5)^;  4. C. Huang 2010a^(6)^;  5. C. Huang 2010b^(7)^;  7. C. Huang 2013^(8)^;  10. C. Huang 2014^(9)^;  14. W. Fan 2015^(10)^;  16. H. Xu 2016^(11)^;  17. L. Zhang 2017^(12)^;  18. H. Xu 2018^(13)^;  19. P. He 2018a^(14)^;  20. P. He 2018b^(15)^;  24. J. Li 2021^(16)^;  26. Z. Cheng 2021a^(17)^;  27. Z. Cheng 2021b^(18)^;  28. P. Sur 2021^(19)^;  29. Y. Li 2021^(20)^;  30. Z. Cheng 2021c^(21)^;  31. D. Xie 2022^(22)^ | 1% China 1990 census | Average cohort size | Province level, prefecture level, and county level | 1959-61 | 1956-58 & 1962-64 |
| 6. Q. Li 2012^(23)^ | A Compilation of Population Statistical Data of Shandong 1949–1984. (No exact census reported) | Average cohort size | County level | 1959-61 | 1956-58 & 1962-64 |
| 21. H. Fang 2019^(24)^ | 1% China 1990 census | Average cohort size | County level | 1959-61 | 1954-57 |
| 22. Q. Cheng 2020^(25)^ | 2000 population census of Sichuan Province | Average cohort size | Prefecture level | 1958-62 | 1953-57 & 1963-67 |
| 8. S. Song 2013a^(26)^;  9. S. Song 2013b^(27)^;  12. S. Song 2014^(28)^;  24. Z. Cheng 2021b^(18)^ | 1% China 1982 census | Multiple imputation algorithm | Province level | 1959-61 | 1951-58 & 1962-70 |
| 11. A. Garnaut 2014^(29)^ | 1% China 2000 census | Linear trend | County level | 1957-60 | 1952-54 & 1963-65 |
| 13. Y. Chen 2015^(30)^ | 1% China 2000 census | Linear trend | County level | 1958-61 | 1952-54 & 63-65 |
| 15. J. Chu 2016^(31)^ | 1% China 1990 census | Linear trend | County level | 1959-61 | 1949-57 & 1963-70 |

#### Table A2. Province level observed population in 1959-61, projected population in 1959-61 based on either pre-famine births (1950-1957) or pre- and post-famine births combined (1950-57 & 1963-1970), and their difference.

| Province | Observed  population  in 1959-61 | Projected  population  in 1959-61^a^ | |  | Difference | |
| --- | --- | --- | --- | --- | --- | --- |
|  |  | **Pre** | **Pre & post** |  | **Pre** | **Pre & post** |
| Sichuan (SC) | 21,938 | 52,806 | 60,429 |  | 30,868 | 38,491 |
| Anhui (AH) | 11,656 | 22,007 | 30,428 |  | 10,351 | 18,772 |
| Guizhou (GZ) | 6,827 | 13,652 | 15,443 |  | 6,825 | 8,616 |
| Hunan (HuN) | 15,157 | 30,479 | 32,945 |  | 15,322 | 17,788 |
| Henan (HeN) | 24,450 | 40,468 | 46,983 |  | 16,018 | 22,533 |
| Qinghai (QH) | 1,025 | 1,587 | 2,119 |  | 562 | 1,094 |
| Jiangsu (JS) | 23,222 | 42,083 | 39,186 |  | 18,861 | 15,964 |
| Ningxia (NX) | 1,161 | 1,835 | 2,264 |  | 674 | 1,103 |
| Guangxi (GX) | 10,967 | 18,427 | 19,901 |  | 7,460 | 8,934 |
| Gansu (GS) | 6,664 | 10,243 | 12,758 |  | 3,579 | 6,094 |
| Shandong (SD) | 30,878 | 52,916 | 50,721 |  | 22,038 | 19,843 |
| Yunnan (YN) | 10,837 | 17,807 | 18,403 |  | 6,970 | 7,566 |
| Hubei (HuB) | 17,484 | 30,623 | 27,740 |  | 13,139 | 10,256 |
| Fujian (FJ) | 9,852 | 16,119 | 14,846 |  | 6,267 | 4,994 |
| Hebei (HeB) | 23,406 | 37,512 | 35,279 |  | 14,106 | 11,873 |
| Zhejiang (ZJ) | 17,087 | 27,955 | 24,105 |  | 10,868 | 7,018 |
| Jiangxi (JX) | 12,153 | 18,438 | 17,860 |  | 6,285 | 5,707 |
| Guangdong (GD) | 23,293 | 36,064 | 32,807 |  | 12,771 | 9,514 |
| Liaoning (LN) | 18,324 | 29,553 | 24,090 |  | 11,229 | 5,766 |
| Shanxi (SX) | 12,059 | 16,667 | 15,408 |  | 4,608 | 3,349 |
| Shaanxi (SaX) | 13,586 | 17,747 | 16,841 |  | 4,161 | 3,255 |
| Jilin (JL) | 11,835 | 14,983 | 14,710 |  | 3,148 | 2,875 |
| Inner Mongolia (IM) | 9,532 | 12,382 | 11,272 |  | 2,850 | 1,740 |
| Heilongjiang (HLJ) | 15,529 | 18,731 | 18,037 |  | 3,202 | 2,508 |
| Beijing (BJ) | 5,594 | 7,762 | 5,317 |  | 2,168 | -277 |
| Tianjin (TJ) | 4,293 | 6,554 | 4,804 |  | 2,261 | 511 |
| Shanghai (SH) | 7,487 | 12,173 | 7,427 |  | 4,686 | -60 |
| Xinjiang (XJ) | 4,949 | 3,566 | 6,328 |  | -1,383 | 1,379 |
| Tibet (TB) | 863 | 641 | 882 |  | -222 | 19 |

*Notes:* ^a^ Projected cohort size estimated from linear regression either based on either pre-famine births (1950-1957) or pre- & post-famine births combined (1950-1957 & 1963-1970) at the province level.

Difference: Projected population – Observed population

*Source:* 1% China 2000 Census, place of birth.

#### Table A3. P-CSSIs, pp-CSSIs, difference and average.

|  | P-CSSI | |  | Pp-CSSI | |  | Difference | |  | Average of ‘P-CSSI’ and ‘Pp-CSSI’ | |
| --- | --- | --- | --- | --- | --- | --- | --- | --- | --- | --- | --- |
| Province | Value | Rank |  | Value | Rank |  | Value | Rank |  | Value | Rank |
| Sichuan (SC) | 58.5 | 1 |  | 63.7 | 1 |  | -5.2 | 18 |  | 61.1 | 1 |
| Anhui (AH) | 47.0 | 4 |  | 61.7 | 2 |  | -14.7 | 23 |  | 54.4 | 2 |
| Guizhou (GZ) | 50.0 | 3 |  | 55.8 | 3 |  | -5.8 | 19 |  | 52.9 | 3 |
| Hunan (HuN) | 50.3 | 2 |  | 54.0 | 4 |  | -3.7 | 16 |  | 52.1 | 4 |
| Henan (HeN) | 39.6 | 9 |  | 48.0 | 7 |  | -8.4 | 20 |  | 43.8 | 5 |
| Qinghai (QH) | 35.4 | 16 |  | 51.6 | 5 |  | -16.2 | 24 |  | 43.5 | 6 |
| Jiangsu (JS) | 44.8 | 5 |  | 40.7 | 11 |  | 4.1 | 9 |  | 42.8 | 7 |
| Ningxia (NX) | 36.7 | 15 |  | 48.7 | 6 |  | -12.0 | 21 |  | 42.7 | 8 |
| Guangxi (GX) | 40.5 | 8 |  | 44.9 | 9 |  | -4.4 | 17 |  | 42.7 | 9 |
| Gansu (GS) | 34.9 | 18 |  | 47.8 | 8 |  | -12.8 | 22 |  | 41.4 | 10 |
| Shandong (SD) | 41.6 | 7 |  | 39.1 | 12 |  | 2.5 | 12 |  | 40.4 | 11 |
| Yunnan (YN) | 39.1 | 10 |  | 41.1 | 10 |  | -2.0 | 15 |  | 40.1 | 12 |
| Hubei (HuB) | 42.9 | 6 |  | 37.0 | 13 |  | 5.9 | 5 |  | 39.9 | 13 |
| Fujian (FJ) | 38.9 | 11 |  | 33.6 | 15 |  | 5.2 | 7 |  | 36.3 | 14 |
| Hebei (HeB) | 37.6 | 14 |  | 33.7 | 14 |  | 4.0 | 10 |  | 35.6 | 15 |
| Zhejiang (ZJ) | 38.9 | 12 |  | 29.1 | 17 |  | 9.8 | 2 |  | 34.0 | 16 |
| Jiangxi (JX) | 34.1 | 19 |  | 32.0 | 16 |  | 2.1 | 13 |  | 33.0 | 17 |
| Guangdong (GD) | 35.4 | 17 |  | 29.0 | 18 |  | 6.4 | 4 |  | 32.2 | 18 |
| Liaoning (LN) | 38.0 | 13 |  | 23.9 | 19 |  | 14.1 | 1 |  | 31.0 | 19 |
| Shanxi (SX) | 27.6 | 20 |  | 21.7 | 20 |  | 5.9 | 6 |  | 24.7 | 20 |
| Shaanxi (SaX) | 23.4 | 21 |  | 19.3 | 22 |  | 4.1 | 8 |  | 21.4 | 21 |
| Jilin (JL) | 21.0 | 23 |  | 19.5 | 21 |  | 1.5 | 14 |  | 20.3 | 22 |
| Inner Mongolia (IM) | 23.0 | 22 |  | 15.4 | 23 |  | 7.6 | 3 |  | 19.2 | 23 |
| Heilongjiang (HLJ) | 17.1 | 24 |  | 13.9 | 24 |  | 3.2 | 11 |  | 15.5 | 24 |

*Notes:* CSSI (%): (Projected population – Observed population) ×100/Projected population

P-CSSI: The projected population is based on pre-famine births (1950-57) in Table A2.

Pp-CSSI: The projected population is based on pre- and post-famine births (1950-57 & 1963-70) in Table A2.

Difference: p-CSSI – pp-CSSI

Provinces are ordered by the average of p-CSSIs and pp-CSSIs from high to low.

*Source:* 1% China 2000 Census, place of birth.

#### Table A4. P-CSSIs, pp-CSSIs, difference and average for five special regions

|  | P-CSSI | Pp-CSSI | Difference | Average of ‘P-CSSI’ and  ‘Pp-CSSI’ |
| --- | --- | --- | --- | --- |
| Beijing (BJ) | 27.9 | -5.2 | 33.1 | 11.4 |
| Tianjin (TJ) | 34.5 | 10.6 | 23.9 | 22.6 |
| Shanghai (SH) | 38.5 | -0.8 | 39.3 | 18.8 |
| Xinjiang (XJ) | -38.8 | 21.8 | -60.6 | -8.5 |
| Tibet (TB) | -34.6 | 2.1 | -36.7 | -16.2 |

*Notes:* CSSI (%): (Projected population – Observed population) × 100/Projected population

P-CSSI: The projected population is based on pre-famine births (1950-57) in Table A2.

Pp-CSSI: The projected population is based on pre- and post-famine births (1950-57 & 1963-70) in Table A2.

Difference: p-CSSI – pp-CSSI

*Source:* 1% China 2000 Census, place of birth.

#### Table A5. Between p-CSSIs correlation at the province level by data source ^a^

|  | 2000  place of birth | 2000  place of residence | 1990  place of residence |
| --- | --- | --- | --- |
| 2000  place of birth | 1 |  |  |
| 2000  place of residence | 0.9795*** | 1 |  |
| 1990  place of residence | 0.9682*** | 0.9518*** | 1 |

*Notes:* 2000 place of birth: CSSI (%) based on place of birth from 1% China 2000 Census.

2000 place of residence: CSSI (%) based on place of residence from 1% China 2000 Census.

1990 place of residence: CSSI (%) based on place of residence from 1% China 1990 Census.

^a^ Pearson’s correlation was used.

****p* < 0.001

#### Table A6. Distribution of p-CSSIs at the prefecture level for each region

| Region | P-CSSI (%)^a^ | | | | | |
| --- | --- | --- | --- | --- | --- | --- |
|  | **N** | **Mean** | **SD** | **Min** | **Max** | **Range** |
| Sichuan (SC) | 22 | 56.8 | 9.6 | 26.3 | 68.7 | 42.4 |
| Anhui (AH) | 17 | 45.1 | 26.8 | -16.9 | 71.6 | 88.5 |
| Guizhou (GZ) | 9 | 49.8 | 4.0 | 46.5 | 58.8 | 12.3 |
| Hunan (HuN) | 14 | 51.9 | 7.8 | 40.6 | 66.0 | 25.5 |
| Henan (HeN) | 17 | 39.8 | 7.5 | 23.4 | 52.8 | 29.4 |
| Qinghai (QH) | 8 | 34.5 | 13.7 | 4.9 | 45.6 | 40.7 |
| Jiangsu (JS) | 13 | 44.5 | 6.0 | 32.6 | 54.8 | 22.2 |
| Ningxia (NX) | 4 | 38.7 | 2.9 | 35.3 | 42.0 | 6.7 |
| Guangxi (GX) | 14 | 38.9 | 7.8 | 27.8 | 55.0 | 27.2 |
| Gansu (GS) | 14 | 34.8 | 9.9 | 7.3 | 48.5 | 41.2 |
| Shandong (SD) | 17 | 41.0 | 11.5 | 19.7 | 65.0 | 45.3 |
| Yunnan (YN) | 16 | 36.3 | 16.1 | -1.4 | 55.4 | 56.8 |
| Hubei (HuB) | 14 | 42.9 | 5.0 | 34.2 | 52.2 | 18.0 |
| Fujian (FJ) | 9 | 38.1 | 5.7 | 29.6 | 45.6 | 15.9 |
| Hebei (HeB) | 11 | 35.9 | 3.5 | 30.0 | 40.8 | 10.9 |
| Zhejiang (ZJ) | 11 | 37.8 | 4.8 | 24.7 | 43.6 | 18.8 |
| Jiangxi (JX) | 11 | 35.2 | 7.9 | 18.4 | 45.5 | 27.2 |
| Guangdong (GD) | 24 | 34.0 | 8.1 | 11.2 | 46.8 | 35.6 |
| Liaoning (LN) | 14 | 35.7 | 6.0 | 24.3 | 44.5 | 20.2 |
| Shanxi (SX) | 11 | 28.8 | 7.8 | 16.1 | 42.5 | 26.4 |
| Shaanxi (SaX) | 10 | 22.6 | 8.5 | 8.8 | 39.3 | 30.6 |
| Jilin (JL) | 9 | 22.7 | 5.6 | 15.4 | 31.9 | 16.5 |
| Inner Mongolia (IM) | 12 | 27.6 | 7.3 | 18.2 | 44.1 | 25.9 |
| Heilongjiang (HLJ) | 13 | 19.3 | 9.8 | -1.7 | 31.8 | 33.5 |
| Xinjiang (XJ) | 16 | -3.3 | 59.5 | -147.6 | 47.3 | 194.9 |
| Tibet (TB) | 7 | -32.7 | 32.8 | -81.4 | 7.4 | 88.8 |
| Beijing (BJ) | 1 | 28.1 | 28.1 | 28.1 | 28.1 | NA |
| Shanghai (SH) | 1 | 39.6 | 39.6 | 39.6 | 39.6 | NA |
| Tianjin (TJ) | 1 | 37.4 | 37.4 | 37.4 | 37.4 | NA |
| All | 340 | 34.7 | 22.8 | -147.6 | 71.6 | 219.2 |

*Notes:* ^a^ Distribution summary statistics of CSSI at the prefecture level for each province generated from Table A7

NA: not applicable

*Source:* 1% China 2000 Census, place of residence.

#### Table A7. CSSIs by prefecture of residence (N=340), 1% China 2000 Census.

| Prefecture at residence | Code | P-CSSI | Pp-CSSI | Average of ‘p-CSSI’ and  ‘pp-CSSI’ |
| --- | --- | --- | --- | --- |
| Sichuan (SC) |  |  |  |  |
| Chongqing municipality | 500000 | 58.9 | 57.1 | 58.0 |
| Chengdu city | 510100 | 55.1 | 58.9 | 57.0 |
| Zigong city | 510300 | 61.8 | 56.9 | 59.4 |
| Panzhihua city | 510400 | 26.3 | 53.9 | 40.1 |
| Luzhou city | 510500 | 61.7 | 57.6 | 59.7 |
| Deyang city | 510600 | 61.1 | 62.0 | 61.5 |
| Mianyang city | 510700 | 54.5 | 55.8 | 55.1 |
| Guangyuan city | 510800 | 60.0 | 56.3 | 58.1 |
| Suining city | 510900 | 55.6 | 57.5 | 56.5 |
| Neijiang city | 511000 | 56.2 | 57.1 | 56.6 |
| Leshan city | 511100 | 68.7 | 65.6 | 67.2 |
| Nanchong city | 511300 | 67.8 | 61.5 | 64.6 |
| Meishan city | 511400 | 68.7 | 68.1 | 68.4 |
| Yibin city | 511500 | 64.5 | 59.0 | 61.8 |
| Guang'an city | 511600 | 53.8 | 52.8 | 53.3 |
| Dazhou city | 511700 | 56.9 | 58.2 | 57.6 |
| Ya'an city | 511800 | 63.1 | 64.8 | 63.9 |
| Bazhong city | 511900 | 61.8 | 57.8 | 59.8 |
| Ziyang city | 512000 | 55.6 | 61.7 | 58.7 |
| Ngawa Tibetan-Qiang autonomous prefecture | 513200 | 48.4 | 48.9 | 48.7 |
| Garze Tibetan autonomous prefecture | 513300 | 38.8 | 36.7 | 37.7 |
| Liangshan Yi prefecture | 513400 | 48.0 | 52.7 | 50.3 |
| Anhui (AH) |  |  |  |  |
| Hefei city | 340100 | 57.6 | 62.4 | 60.0 |
| Wuhu city | 340200 | 64.8 | 62.9 | 63.9 |
| Bengbu city | 340300 | 37.6 | 46.5 | 42.1 |
| Huainan city | 340400 | 46.9 | 46.3 | 46.6 |
| Ma'anshan city | 340500 | 57.2 | 57.2 | 57.2 |
| Huaibei city | 340600 | 3.7 | 46.4 | 25.0 |
| Tongling city | 340700 | 46.4 | 51.9 | 49.1 |
| Anqing city | 340800 | 61.3 | 60.5 | 60.9 |
| Huangshan city | 341000 | 66.8 | 61.7 | 64.2 |
| Chuzhou city | 341100 | 56.6 | 61.0 | 58.8 |
| Fuyang city | 341200 | -1.6 | 41.5 | 19.9 |
| Suzhou city | 341300 | 26.7 | 52.0 | 39.3 |
| Chaohu city | 341400 | 68.4 | 72.7 | 70.6 |
| Liu'an city | 341500 | 53.7 | 60.5 | 57.1 |
| Bozhou city | 341600 | -16.9 | 43.2 | 13.1 |
| Guichi city | 341700 | 66.1 | 59.3 | 62.7 |
| Xuancheng city | 341800 | 71.6 | 72.9 | 72.3 |
| Guizhou (GZ) |  |  |  |  |
| Guiyang city | 520100 | 46.8 | 40.8 | 43.8 |
| Liupanshui city | 520200 | 46.5 | 39.9 | 43.2 |
| Zunyi city | 520300 | 48.5 | 60.7 | 54.6 |
| Anshun city | 520400 | 48.6 | 44.1 | 46.4 |
| Tongren prefecture | 522200 | 47.6 | 54.6 | 51.1 |
| Qianxinan Buyei-Miao autonomous prefecture | 522300 | 48.2 | 38.0 | 43.1 |
| Bijie prefecture | 522400 | 58.8 | 50.7 | 54.8 |
| Qiandongnan Miao-Dong autonomous prefecture | 522600 | 49.1 | 52.9 | 51.0 |
| Qiannan Buyei-Miao autonomous prefecture | 522700 | 54.0 | 49.8 | 51.9 |
| Hunan (HuN) |  |  |  |  |
| Changsha city | 430100 | 40.6 | 41.2 | 40.9 |
| Zhuzhou city | 430200 | 51.6 | 46.6 | 49.1 |
| Xiangtan city | 430300 | 47.8 | 53.8 | 50.8 |
| Hengyang city | 430400 | 55.4 | 48.3 | 51.8 |
| Shaoyang city | 430500 | 48.7 | 47.5 | 48.1 |
| Yueyang city | 430600 | 46.1 | 45.1 | 45.6 |
| Changde city | 430700 | 43.9 | 46.6 | 45.3 |
| Zhangjiajie city | 430800 | 66.0 | 67.3 | 66.7 |
| Yiyang city | 430900 | 46.5 | 48.3 | 47.4 |
| Chenzhou city | 431000 | 44.8 | 36.6 | 40.7 |
| Yongzhou city | 431100 | 61.0 | 53.6 | 57.3 |
| Huaihua city | 431200 | 52.8 | 54.0 | 53.4 |
| Loudi city | 431300 | 59.3 | 57.2 | 58.3 |
| Xiangxi Tujia-Miao autonomous prefecture | 433100 | 62.0 | 62.7 | 62.3 |
| Henan (HeN) |  |  |  |  |
| Zhengzhou city | 410100 | 39.5 | 35.9 | 37.7 |
| Kaifeng city | 410200 | 49.6 | 42.4 | 46.0 |
| Luoyang city | 410300 | 42.3 | 37.8 | 40.0 |
| Pingdingshan city | 410400 | 41.2 | 40.5 | 40.8 |
| Anyang city | 410500 | 52.8 | 49.9 | 51.4 |
| Hebi city | 410600 | 41.3 | 43.4 | 42.4 |
| Xinxiang city | 410700 | 42.8 | 41.5 | 42.2 |
| Jiaozuo city | 410800 | 42.9 | 34.5 | 38.7 |
| Puyang city | 410900 | 51.6 | 49.1 | 50.3 |
| Xuchang city | 411000 | 41.8 | 45.6 | 43.7 |
| Luohe city | 411100 | 37.3 | 43.8 | 40.6 |
| Sanmenxia city | 411200 | 31.4 | 27.7 | 29.6 |
| Nanyang city | 411300 | 34.5 | 43.5 | 39.0 |
| Shangqiu city | 411400 | 39.1 | 47.3 | 43.2 |
| Xinyang city | 411500 | 27.8 | 44.4 | 36.1 |
| Zhoukou city | 411600 | 37.3 | 44.7 | 41.0 |
| Zhumadian city | 411700 | 23.4 | 46.7 | 35.1 |
| Qinghai (QH) |  |  |  |  |
| Xining city | 630100 | 45.6 | 47.4 | 46.5 |
| Haidong prefecture | 632100 | 39.9 | 47.8 | 43.9 |
| Haibei Tibetan autonomous prefecture | 632200 | 39.6 | 50.8 | 45.2 |
| Huangnan Tibetan autonomous prefecture | 632300 | 42.9 | 50.4 | 46.7 |
| Hainan Tibetan autonomous prefecture | 632500 | 45.1 | 47.6 | 46.3 |
| Golog Tibetan autonomous prefecture | 632600 | 25.9 | 45.8 | 35.9 |
| Yushu Tibetan autonomous prefecture | 632700 | 4.9 | 49.8 | 27.4 |
| Haixi Mongol-Tibetan autonomous prefecture | 632800 | 32.0 | 48.0 | 40.0 |
| Jiangsu (JS) |  |  |  |  |
| Nanjing city | 320100 | 42.9 | 31.5 | 37.2 |
| Wuxi city | 320200 | 41.1 | 38.4 | 39.8 |
| Xuzhou city | 320300 | 41.0 | 38.7 | 39.9 |
| Changzhou city | 320400 | 32.6 | 34.3 | 33.5 |
| Suzhou city | 320500 | 51.3 | 45.0 | 48.2 |
| Nantong city | 320600 | 43.0 | 34.2 | 38.6 |
| Lianyungang city | 320700 | 44.0 | 31.0 | 37.5 |
| Huaiyin city | 320800 | 49.2 | 35.3 | 42.3 |
| Yancheng city | 320900 | 43.1 | 30.6 | 36.9 |
| Yangzhou city | 321000 | 54.8 | 51.2 | 53.0 |
| Zhenjiang city | 321100 | 46.0 | 42.7 | 44.3 |
| Taizhou city | 321200 | 51.3 | 47.8 | 49.6 |
| Suqian city | 321300 | 38.2 | 30.2 | 34.2 |
| Ningxia (NX) |  |  |  |  |
| Yinchuan city | 640100 | 42.0 | 42.4 | 42.2 |
| Shizuishan city | 640200 | 37.9 | 40.4 | 39.2 |
| Wuzhong city | 640300 | 35.3 | 40.7 | 38.0 |
| Guyuan prefecture | 642200 | 41.4 | 48.3 | 44.8 |
| Guangxi (GX) |  |  |  |  |
| Nanning city | 450100 | 33.4 | 31.3 | 32.4 |
| Liuzhou city | 450200 | 27.8 | 33.2 | 30.5 |
| Guilin city | 450300 | 55.0 | 49.2 | 52.1 |
| Wuzhou city | 450400 | 47.0 | 47.4 | 47.2 |
| Beihai city | 450500 | 35.3 | 28.1 | 31.7 |
| Fangchenggang city | 450600 | 29.4 | 32.4 | 30.9 |
| Qinzhou city | 450700 | 31.7 | 27.9 | 29.8 |
| Guigang city | 450800 | 43.9 | 43.6 | 43.7 |
| Yulin city | 450900 | 46.7 | 41.9 | 44.3 |
| Nanning prefecture | 452100 | 34.0 | 39.8 | 36.9 |
| Liuzhou prefecture | 452200 | 45.2 | 47.2 | 46.2 |
| Hezhou prefecture | 452400 | 31.0 | 44.2 | 37.6 |
| Baise prefecture | 452600 | 46.3 | 44.1 | 45.2 |
| Hechi prefecture | 452700 | 34.6 | 38.1 | 36.3 |
| Gansu (GS) |  |  |  |  |
| Lanzhou city | 620100 | 35.3 | 33.8 | 34.6 |
| Jiayuguan city | 620200 | 46.0 | 49.6 | 47.8 |
| Jinchang city | 620300 | 34.7 | 51.5 | 43.1 |
| Baiyin city | 620400 | 40.6 | 39.8 | 40.2 |
| Tianshui city | 620500 | 39.9 | 47.9 | 43.9 |
| Jiuquan prefecture | 622100 | 41.7 | 45.1 | 43.4 |
| Zhangye prefecture | 622200 | 33.4 | 43.3 | 38.3 |
| Wuwei prefecture | 622300 | 35.1 | 39.8 | 37.4 |
| Dingxi prefecture | 622400 | 30.0 | 44.2 | 37.1 |
| Wudu prefecture | 622600 | 35.2 | 50.5 | 42.8 |
| Pingliang prefecture | 622700 | 32.4 | 47.2 | 39.8 |
| Qingyang prefecture | 622800 | 27.4 | 35.0 | 31.2 |
| Linxia Hui prefecture | 622900 | 7.3 | 36.0 | 21.7 |
| Gannan Tibetan autonomous prefecture | 623000 | 48.5 | 50.6 | 49.5 |
| Shandong (SD) |  |  |  |  |
| Jinan city | 370100 | 39.7 | 29.9 | 34.8 |
| Qingdao city | 370200 | 50.9 | 40.1 | 45.5 |
| Zibo city | 370300 | 43.5 | 32.3 | 37.9 |
| Zaozhuang city | 370400 | 37.6 | 32.5 | 35.1 |
| Dongying city | 370500 | 61.1 | 58.7 | 59.9 |
| Yantai city | 370600 | 37.4 | 24.6 | 31.0 |
| Weifang city | 370700 | 40.9 | 33.2 | 37.1 |
| Jining city | 370800 | 37.7 | 34.2 | 36.0 |
| Tai'an prefecture | 370900 | 19.7 | 21.0 | 20.3 |
| Weihai city | 371000 | 44.8 | 27.1 | 36.0 |
| Rizhao city | 371100 | 34.2 | 32.6 | 33.4 |
| Laiwu city | 371200 | 31.7 | 20.4 | 26.0 |
| Linyi city | 371300 | 27.3 | 27.0 | 27.1 |
| Dezhou city | 371400 | 53.1 | 48.2 | 50.6 |
| Liaocheng city | 371500 | 37.3 | 37.6 | 37.4 |
| Binzhou city | 371600 | 65.0 | 55.9 | 60.5 |
| Heze city | 371700 | 34.4 | 45.7 | 40.0 |
| Yunnan (YN) |  |  |  |  |
| Kunming city | 530100 | 40.1 | 38.5 | 39.3 |
| Qujing city | 530300 | 31.7 | 33.1 | 32.4 |
| Yuxi city | 530400 | 55.4 | 53.7 | 54.5 |
| Zhaotong prefecture | 532100 | 41.3 | 36.8 | 39.1 |
| Chuxiong Yi autonomous autonomous | 532300 | 48.9 | 45.4 | 47.1 |
| Honghe Hani-Yi autonomous prefecture | 532500 | 41.0 | 38.0 | 39.5 |
| Wenshan Zhuang-Miao autonomous prefecture | 532600 | 40.1 | 39.4 | 39.7 |
| Simao prefecture | 532700 | 38.3 | 30.9 | 34.6 |
| Xishuangpanna Dai autonomous prefecture | 532800 | 8.5 | 20.5 | 14.5 |
| Dali Bai autonomous prefecture | 532900 | 45.7 | 46.4 | 46.1 |
| Baoshan prefecture | 533000 | 46.0 | 35.7 | 40.9 |
| Dehong Dai-Jingpo autonomous prefecture | 533100 | 10.9 | 7.1 | 9.0 |
| Lijiang prefecture | 533200 | 44.8 | 41.7 | 43.2 |
| Nujiang Lisu autonomous prefecture | 533300 | -1.4 | 20.1 | 9.3 |
| Diqing Tibetan autonomous prefecture | 533400 | 41.6 | 43.2 | 42.4 |
| Lincang prefecture | 533500 | 47.8 | 39.6 | 43.7 |
| Hubei (HuB) |  |  |  |  |
| Wuhan city | 420100 | 40.9 | 26.5 | 33.7 |
| Huangshi city | 420200 | 40.0 | 35.7 | 37.8 |
| Shiyan city | 420300 | 34.2 | 35.7 | 34.9 |
| Yichang city | 420500 | 46.4 | 34.7 | 40.6 |
| Xiangfan city | 420600 | 38.8 | 32.7 | 35.8 |
| Ezhou city | 420700 | 46.1 | 41.0 | 43.5 |
| Jingmen city | 420800 | 38.7 | 29.6 | 34.2 |
| Xiaogan city | 420900 | 47.4 | 33.6 | 40.5 |
| Jingzhou city | 421000 | 41.3 | 30.8 | 36.0 |
| Huanggang city | 421100 | 50.3 | 37.7 | 44.0 |
| Xianning city | 421200 | 42.7 | 28.8 | 35.8 |
| Suizhou city | 421300 | 42.0 | 27.0 | 34.5 |
| Exi Tujia-Miao autonomous prefecture | 422800 | 52.2 | 51.3 | 51.7 |
| Hubei Province direct administrative | 429000 | 44.9 | 33.3 | 39.1 |
| Fujian (FJ) |  |  |  |  |
| Fuzhou city | 350100 | 37.8 | 28.1 | 32.9 |
| Xiamen city | 350200 | 29.6 | 25.3 | 27.5 |
| Putian city | 350300 | 38.5 | 25.8 | 32.1 |
| Sanming city | 350400 | 29.8 | 34.1 | 31.9 |
| Quanzhou city | 350500 | 40.0 | 34.1 | 37.0 |
| Zhangzhou city | 350600 | 35.5 | 27.4 | 31.4 |
| Nanping city | 350700 | 42.2 | 37.8 | 40.0 |
| Longyan city | 350800 | 45.6 | 41.0 | 43.3 |
| Ningde city | 350900 | 44.1 | 28.8 | 36.5 |
| Hebei (HeB) |  |  |  |  |
| Shijiazhuang city | 130100 | 40.8 | 30.7 | 35.8 |
| Tangshan city | 130200 | 38.0 | 28.4 | 33.2 |
| Qinhuangdao city | 130300 | 35.3 | 23.5 | 29.4 |
| Handan city | 130400 | 40.3 | 35.2 | 37.8 |
| Xingtai city | 130500 | 39.6 | 35.4 | 37.5 |
| Baoding city | 130600 | 35.9 | 30.6 | 33.2 |
| Zhangjiakou city | 130700 | 35.1 | 23.6 | 29.4 |
| Chengde city | 130800 | 33.1 | 14.4 | 23.7 |
| Cangzhou city | 130900 | 31.8 | 29.0 | 30.4 |
| Langfang city | 131000 | 30.0 | 30.5 | 30.3 |
| Hengshui city | 131100 | 35.2 | 34.3 | 34.8 |
| Zhejiang (ZJ) |  |  |  |  |
| Hangzhou city | 330100 | 35.8 | 18.3 | 27.1 |
| Ningbo city | 330200 | 37.7 | 27.8 | 32.8 |
| Wenzhou city | 330300 | 39.3 | 31.4 | 35.4 |
| Jiaxing city | 330400 | 43.6 | 33.0 | 38.3 |
| Huzhou city | 330500 | 36.8 | 29.1 | 33.0 |
| Shaoxing city | 330600 | 38.9 | 29.0 | 34.0 |
| Jinhua city | 330700 | 40.9 | 36.9 | 38.9 |
| Quzhou city | 330800 | 38.4 | 26.1 | 32.2 |
| Zhoushan city | 330900 | 24.7 | 9.4 | 17.1 |
| Taizhou city | 331000 | 41.2 | 29.3 | 35.3 |
| Lishui city | 331100 | 38.9 | 30.0 | 34.4 |
| Jiangxi (JX) |  |  |  |  |
| Nanchang city | 360100 | 29.2 | 26.6 | 27.9 |
| Jingdezhen city | 360200 | 18.4 | 14.7 | 16.5 |
| Pingxiang city | 360300 | 43.0 | 30.2 | 36.6 |
| Jiujiang city | 360400 | 42.8 | 33.0 | 37.9 |
| Xinyu city | 360500 | 34.7 | 27.0 | 30.9 |
| Yingtan city | 360600 | 36.8 | 26.4 | 31.6 |
| Ganzhou city | 360700 | 39.2 | 29.2 | 34.2 |
| Ji’an city | 360800 | 34.8 | 23.8 | 29.3 |
| Yichun city | 360900 | 35.8 | 23.9 | 29.9 |
| Fuzhou city | 361000 | 45.5 | 38.5 | 42.0 |
| Shangrao city | 361100 | 27.5 | 21.1 | 24.3 |
| Guangdong (GD) |  | |  |  |
| Guangzhou city | 440100 | 37.0 | 28.8 | 32.9 |
| Shaoguan city | 440200 | 32.3 | 31.3 | 31.8 |
| Shenzhen city | 440300 | 25.3 | 48.6 | 36.9 |
| Zhuhai city | 440400 | 31.9 | 34.2 | 33.1 |
| Shantou city | 440500 | 34.3 | 21.3 | 27.8 |
| Foshan city | 440600 | 33.4 | 41.9 | 37.7 |
| Jiangmen city | 440700 | 37.3 | 26.1 | 31.7 |
| Zhanjiang city | 440800 | 29.3 | 18.4 | 23.8 |
| Maoming city | 440900 | 28.9 | 22.4 | 25.7 |
| Zhaoqing city | 441200 | 41.3 | 37.6 | 39.4 |
| Huizhou city | 441300 | 33.2 | 34.8 | 34.0 |
| Meizhou city | 441400 | 40.4 | 24.8 | 32.6 |
| Shanwei city | 441500 | 44.7 | 31.1 | 37.9 |
| Heyuan city | 441600 | 42.3 | 29.7 | 36.0 |
| Yangjiang city | 441700 | 30.3 | 30.6 | 30.4 |
| Qingyuan city | 441800 | 46.1 | 35.0 | 40.6 |
| Dongguan city | 441900 | 26.4 | 50.4 | 38.4 |
| Zhongshan city | 442000 | 34.4 | 40.7 | 37.5 |
| Chaozhou city | 445100 | 46.8 | 33.7 | 40.3 |
| Jieyang city | 445200 | 38.5 | 24.4 | 31.5 |
| Yunfu city | 445300 | 38.8 | 31.3 | 35.1 |
| Hainan Province direct administrative | 460000 | 26.9 | 26.8 | 26.8 |
| Haikou city | 460100 | 24.2 | 32.1 | 28.1 |
| Sanya city | 460200 | 11.2 | 29.2 | 20.2 |
| Liaoning (LN) |  |  |  |  |
| Shenyang city | 210100 | 38.0 | 18.4 | 28.2 |
| Dalian city | 210200 | 39.0 | 19.6 | 29.3 |
| Anshan city | 210300 | 38.2 | 21.8 | 30.0 |
| Fushun city | 210400 | 24.3 | 10.5 | 17.4 |
| Benxi city | 210500 | 27.7 | 11.0 | 19.3 |
| Dandong city | 210600 | 37.9 | 26.9 | 32.4 |
| Jinzhou city | 210700 | 44.1 | 31.3 | 37.7 |
| Yingkou city | 210800 | 29.9 | 21.5 | 25.7 |
| Fuxin city | 210900 | 36.3 | 20.7 | 28.5 |
| Liaoyang city | 211000 | 40.2 | 27.1 | 33.7 |
| Panjin city | 211100 | 37.3 | 34.4 | 35.9 |
| Tieling city | 211200 | 29.8 | 27.0 | 28.4 |
| Chaoyang city | 211300 | 33.0 | 15.6 | 24.3 |
| Huludao city | 211400 | 44.5 | 28.3 | 36.4 |
| Shanxi (SX) |  |  |  |  |
| Taiyuan city | 140100 | 23.8 | 15.9 | 19.9 |
| Datong city | 140200 | 29.8 | 20.4 | 25.1 |
| Yangquan city | 140300 | 40.7 | 29.7 | 35.2 |
| Changzhi city | 140400 | 29.4 | 22.1 | 25.8 |
| Jincheng city | 140500 | 42.5 | 29.8 | 36.1 |
| Shuozhou city | 140600 | 30.7 | 16.9 | 23.8 |
| Jinzhong city | 140700 | 31.2 | 26.0 | 28.6 |
| Yuncheng city | 140800 | 25.7 | 17.8 | 21.8 |
| Xinzhou city | 140900 | 16.1 | 11.4 | 13.7 |
| Linfen city | 141000 | 20.5 | 13.1 | 16.8 |
| Luliang prefecture | 142300 | 26.9 | 26.0 | 26.5 |
| Shannxi (SaX) |  |  |  |  |
| Xi'an city | 610100 | 27.9 | 15.3 | 21.6 |
| Tongchuan city | 610200 | 20.5 | 13.2 | 16.8 |
| Baoji city | 610300 | 23.7 | 19.4 | 21.6 |
| Xianyang city | 610400 | 21.9 | 19.3 | 20.6 |
| Weinan city | 610500 | 19.4 | 14.5 | 16.9 |
| Yan'an city | 610600 | 8.8 | 10.1 | 9.4 |
| Hanzhong city | 610700 | 30.3 | 25.5 | 27.9 |
| Yulin city | 610800 | 14.1 | 6.6 | 10.3 |
| Ankang city | 610900 | 39.3 | 24.4 | 31.8 |
| Shangluo prefecture | 612500 | 19.8 | 25.1 | 22.5 |
| Jilin (JL) |  |  |  |  |
| Changchun city | 220100 | 20.5 | 14.3 | 17.4 |
| Jilin city | 220200 | 17.5 | 17.1 | 17.3 |
| Siping city | 220300 | 28.7 | 23.0 | 25.9 |
| Liaoyuan city | 220400 | 19.0 | 19.5 | 19.3 |
| Tonghua city | 220500 | 28.3 | 22.0 | 25.1 |
| Baishan city | 220600 | 31.9 | 24.3 | 28.1 |
| Songyuan city | 220700 | 21.1 | 16.9 | 19.0 |
| Baicheng city | 220800 | 15.4 | 12.5 | 14.0 |
| Yanbian Korean autonomous prefecture | 222400 | 21.7 | 10.7 | 16.2 |
| Inner Mongolia (IM) |  | |  |  |
| Hohhot city | 150100 | 18.2 | 13.2 | 15.7 |
| Baotou city | 150200 | 24.5 | 15.7 | 20.1 |
| Wuhai city | 150300 | 31.6 | 19.3 | 25.5 |
| Chifeng city | 150400 | 24.8 | 4.7 | 14.7 |
| Tongliao city | 150500 | 24.8 | 20.5 | 22.7 |
| Hulunbuir league | 152100 | 22.1 | 23.2 | 22.7 |
| Xing'an league | 152200 | 27.7 | 21.8 | 24.7 |
| Xilingol league | 152500 | 21.7 | 12.5 | 17.1 |
| Ulaanchab league | 152600 | 29.5 | 9.5 | 19.5 |
| Yikezhao league | 152700 | 23.7 | 14.9 | 19.3 |
| Bayannur league | 152800 | 44.1 | 31.8 | 38.0 |
| Alxa prefecture | 152900 | 38.0 | 41.0 | 39.5 |
| Heilongjiang (HLJ) |  | |  |  |
| Harbin city | 230100 | 20.5 | 9.9 | 15.2 |
| Qiqihar city | 230200 | 25.9 | 17.4 | 21.6 |
| Jixi city | 230300 | 20.1 | 19.9 | 20.0 |
| Hegang city | 230400 | -1.7 | 8.0 | 3.1 |
| Shuangyashan city | 230500 | 19.2 | 15.1 | 17.2 |
| Daqing city | 230600 | 2.2 | 17.5 | 9.9 |
| Yichun city | 230700 | 12.4 | 10.8 | 11.6 |
| Jiamusi city | 230800 | 28.7 | 17.5 | 23.1 |
| Qitaihe city | 230900 | 23.5 | 12.4 | 18.0 |
| Mudanjiang city | 231000 | 25.6 | 18.8 | 22.2 |
| Heihe city | 231100 | 18.9 | 10.3 | 14.6 |
| Suihua city | 231200 | 23.1 | 12.1 | 17.6 |
| Daxing'anling prefecture | 232700 | 31.8 | 25.9 | 28.9 |
| Xinjiang (XJ) |  |  |  |  |
| Urumuqi city | 650100 | 20.4 | 26.5 | 23.5 |
| Karamay city | 650200 | 40.0 | 37.8 | 38.9 |
| Turfan prefecture | 652100 | 0.2 | 22.6 | 11.4 |
| Hami prefecture | 652200 | 18.1 | 30.1 | 24.1 |
| Changji Hui prefecture | 652300 | 23.5 | 33.3 | 28.4 |
| Bortala Mongol autonomous prefecture | 652700 | 35.1 | 36.6 | 35.8 |
| Bayin'gholin Mongol autonomous prefecture | 652800 | 22.8 | 29.2 | 26.0 |
| Aksu prefecture | 652900 | -53.3 | 25.0 | -14.2 |
| Kizilsu Kirghiz autonomous prefecture | 653000 | -50.1 | 14.2 | -18.0 |
| Kashgar prefecture | 653100 | -147.6 | 13.8 | -66.9 |
| Khotan prefecture | 653200 | -124.3 | 15.6 | -54.4 |
| Yili Kazak autonomous prefecture | 654000 | 47.3 | 50.2 | 48.8 |
| Yili prefecture | 654100 | 12.0 | 30.2 | 21.1 |
| Tacheng prefecture | 654200 | 40.8 | 38.2 | 39.5 |
| Altay prefecture | 654300 | 40.7 | 40.3 | 40.5 |
| Shihezi city | 659000 | 20.9 | 44.1 | 32.5 |
| Tibet (TB) |  |  |  |  |
| Lhasa city | 540100 | 5.5 | 14.0 | 9.7 |
| Chamdo prefecture | 542100 | -81.4 | 8.5 | -36.5 |
| Shannan prefecture | 542200 | -59.2 | -2.5 | -30.9 |
| Shigatse prefecture | 542300 | -22.9 | 4.2 | -9.3 |
| Nagchu prefecture | 542400 | -30.3 | 13.8 | -8.3 |
| Ngari prefecture | 542500 | -47.7 | -4.4 | -26.0 |
| Nyingchi prefecture | 542600 | 7.4 | -0.2 | 3.6 |
| Beijing (BJ) |  |  |  |  |
| Beijing municipality | 110000 | 28.1 | 10.3 | 19.2 |
| Shanghai (SH) |  |  |  |  |
| Shanghai municipality | 310000 | 39.6 | 9.1 | 24.4 |
| Tianjin (TJ) |  |  |  |  |
| Tianjin municipality | 120000 | 37.4 | 14.6 | 26.0 |

*Notes:* CSSI (%): (Projected population – Observed population) × 100/Projected population

P-CSSI: The projected population is based on pre-famine births (1950-57) at prefecture level.

Pp-CSSI: The projected population is based on pre- and post-famine births (1950-57 & 1963-70) at the prefecture level.

*Source:* 1% China 2000 Census, place of residence.

#### Table A8. Distribution of p-CSSIs and pp-CSSIs at the prefecture level within Sichuan Province

| Row # | CSSI calculating method | Years used | Mean | Median | Standard deviation |
| --- | --- | --- | --- | --- | --- |
| 1 | P-CSSI | 1950-57 | 43.4 | 44.8 | 9.7 |
| 2 | Pp-CSSI | 1950-57 & 1963-70 | 50.1 | 50.3 | 6.4 |

#### Table A9. Meta-regression of the association between CSSI and famine cohort TB incidence rate ratio (IRR), tuberculosis surveillance study in Sichuan Province

| Row # | CSSI calculating method | Years used | Estimated log change IRR per unit increase in CSSI (95%CI) | Difference between estimates  (P-CSSI vs. Pp-CSSI)^a^ |
| --- | --- | --- | --- | --- |
| 1 | P-CSSI | 1950-57 | 0.38 (-0.12, 0.88) | 0.55 (0.06, 10.4) |
| 2 | Pp-CSSI | 1950-57 & 1963-70 | 0.91 (0.19, 1.63) |  |

*Notes:* ^a^ Estimate of difference made by bootstrap: bootstrap resampled the original dataset for 1000 times to obtain the estimated variance of the difference of effect estimates for both linear trends in 1950-70 using two datasets. 95% CIs are constructed based on the estimated variance.

### SI References：

1.	X. Meng, N. Qian, The long run health and economic consequences of famine on survivors: Evidence from China's Great Famine. *IZA Discussion Papers*, 2471 (2006).

2.	X. Meng, N. Qian (2009) The long term consequences of famine on survivors: evidence from a unique natural experiment using China's great famine (National Bureau of Economic Research, National Bureau of Economic Research).

18.	Z. Cheng, R. Smyth (2021) Does Childhood Adversity Affect Household Portfolio Decisions? Evidence from the Chinese Great Famine. (ResearchGate, ResearchGate).

19.	P. K. Sur, M. Sasaki, The Persistent Effect of Famine on Present-Day China: Evidence from the Billionaires. *IZA Discussion Papers*, dp14291 (2021).

20.	Y. Li, N. Sunder, What doesn’t kill her, will make her depressed. *Economics & Human Biology* **43**, 101064 (2021).

21.	Z. Cheng, L. Guo, R. Smyth, M. Tani, Childhood Adversity and Energy Poverty. *IZA Discussion Papers*, dp14809 (2021).

22.	D. Xie, Z. Zhu, Intergenerational effects of early-life health shocks during the Chinese 1959–1961 famine. *Ageing and Society*, 1-16 (2022).

23.	Q. Da Li *et al.*, Nutrition deficiency increases the risk of stomach cancer mortality. *BMC Cancer* **12**, 1-12 (2012).

24.	H. Fang, L. Hou, M. Liu, L. C. Xu, P. R. Zhang, Factions, Local Accountability, and Long-Term Development: County-Level Evidence from a Chinese Province (working paper). *National Bureau of Economic Research*, w25901 (2019).

30.	Y. Chen, D. Yang (2015) Historical traumas and the roots of political distrust: Political inference from the Great Chinese Famine. (Social Science Research Network, Social Science Research Network).

31.	J. Chu, I. P. Png, J. Yi (2016) Entrepreneurship and the school of hard knocks: evidence from China's great famine. (Social Science Research Network, Social Science Research Network).
